# Quantum-Refined Latent Diffusion: A Hybrid Generative Framework for Imbalanced ECG Classification

**DOI:** 10.64898/2026.04.09.26350502

**Authors:** Georgios Kritopoulos, Georgios Neofotistos, Georgios D. Barmparis, Giorgos P. Tsironis

## Abstract

Class imbalance in clinical electrocardiogram (ECG) datasets limits the diagnostic sensitivity of automated arrhythmia classifiers, particularly for rare but clinically significant beat types. We propose a three-stage hybrid generative pipeline that combines a spectral-guided conditional Variational Autoencoder (cVAE), a class-conditional latent Denoising Diffusion Probabilistic Model (DDPM), and a Quantum Latent Refinement (QLR) module built on parameterized quantum circuits to augment minority arrhythmia classes in the MIT-BIH Arrhythmia Database. The QLR module applies a bounded residual correction guided by Maximum Mean Discrepancy minimization to align synthetic latent distributions with real class-specific latent banks. A lightweight 1D MobileNetV2 classifier evaluated over five independent random seeds and four augmentation ratios serves as the downstream benchmark. Our findings establish latent diffusion augmentation as an effective strategy for imbalanced ECG classification and motivate further investigation of quantum-classical hybrid methods in cardiac diagnostics.

## 1 Introduction

Cardiovascular diseases remain among the leading causes of mortality, and the electrocardiogram (ECG) is the principal non-invasive tool for their diagnosis. ^1^ By capturing the electrical activity of the heart as a time-series signal, the ECG encodes critical information about cardiac rhythm, conduction velocity, and myocardial repolarization, ^2^ making it essential for identifying arrhythmias in both hospital and ambulatory settings. ^3^ Deep learning approaches - particularly convolutional neural networks (CNNs)^4,5^ and recurrent architectures^6,7^ - have demonstrated competitive performance on automated arrhythmia detection benchmarks. ^3^ The proliferation of wearable cardiac monitors capable of continuous ECG acquisition, generating high volumes of unlabeled data, has further elevated the need for computationally efficient classifiers that are sensitive enough to detect rare pathological events in real time. ^8^

Despite these advances, a fundamental problem remains: class imbalance. In physiological ECG datasets such as the MIT-BIH Arrhythmia Database, ^9^ Normal sinus rhythm beats account for the overwhelming majority of all recorded beats, while clinically significant minority classes Supraventricular ectopic beats, Ventricular ectopic beats, and Fusion beats – represent only a small fraction of the total (see Section 2.1 for a detailed description of beat classes). When standard classifiers undergo training on such distributions, they optimize global accuracy at the expense of per-class sensitivity. This phenomenon, known as the accuracy paradox, ^10^ has been documented across medical machine learning applications: a model may exceed 94% test accuracy while correctly detecting only a small proportion of rare but life-threatening arrhythmia events. From a clinical standpoint this trade-off is unacceptable. Macro F1, which computes the unweighted mean of per-class F1 scores, is therefore adopted as the primary evaluation metric, as it penalizes class-selective failure regardless of class frequency. ^11,12^ The clinical stakes are asymmetric. A missed ventricular ectopic beat may leave a patient at unrecognized risk for progression to ventricular tachycardia or fibrillation, ^13^ whereas excessive false alarms from a low-precision classifier generate alert fatigue that progressively erodes clinician trust in automated monitoring systems. ^14^

Classical oversampling methods such as the Synthetic Minority Oversampling Technique (SMOTE) ^15^ partially address imbalance by generating interpolated minority samples, but linear interpolation in a high-dimensional signal space produces morphologically unrealistic beats that distort the training distribution and tend to degrade performance at high augmentation volumes. ^16^ Generative Adversarial Networks (GANs) offer a more principled approach, but suffer from training instabilities – including mode collapse and blurry reconstructions – that limit their reliability in medical signal synthesis. ^17^ Denoising diffusion probabilistic models (DDPMs) ^18^ avoid many of these issues and have shown strong results in sequential data generation, but their computational demands at the raw signal level can be substantial for large-scale ECG databases. ^19,20^

Quantum Machine Learning (QML) has emerged as a distinct computational paradigm that operates in an exponentially large Hilbert space, potentially implementing feature maps with structured inductive biases that differ from those of classical networks. ^21^ Whether near-term quantum circuits offer practical computational advantages over classical alternatives remains an open and actively debated question – known challenges include barren plateau phenomena (the vanishing of parameter gradients in wide or deep quantum circuits^22^), concentration-of-measure effects, and the classical simulability of many shallow Parameterized Quantum Circuit (PQC) architectures – but hybrid quantum-classical designs provide a pragmatic route for exploring these properties under current hardware constraints.^23,24^ In cardiac diagnostics, quantum-enhanced models have shown early exploratory results for arrhythmia classification^25–28^ and for the generative augmentation of ECG signals,^29^ motivating further investigation of the architectural approach even as its computational advantages remain to be established.^30,31^

This work proposes and evaluates a three-stage hybrid generative pipeline for imbalanced ECG arrhythmia classification. A spectral-guided cVAE first learns a compact latent representation of ECG beats. A class-conditional latent DDPM then synthesizes new minority-class samples in that latent space, and a Quantum Latent Refinement (QLR) module applies a distributional correction to align synthetic latents with the real class-specific manifold. A lightweight 1D MobileNetV2^32^ classifier is trained on the augmented dataset and benchmarked against an unaugmented baseline, SMOTE, cVAE-only, and plain latent DDPM augmentation across five random seeds and four augmentation ratios.

## 2 Materials and Methods

The proposed pipeline consists of three sequential generative stages, each building on the output of the previous one. First, a spectral-guided conditional variational autoencoder (cVAE) learns a compact 32-dimensional latent representation of ECG beats. Second, a class-conditional denoising diffusion probabilistic model (DDPM) is trained on these latent vectors to generate new synthetic minority-class samples. Third, a Quantum Latent Refinement (QLR) module – built on a parameterized quantum circuit – applies a bounded distributional correction to align the DDPM-generated latents with the real class-specific latent manifold. A lightweight 1D MobileNetV2 classifier is then trained on the real beats augmented by synthetic ones and evaluated using Macro F1. Each component is described in detail in Sections 2.3–2.6. An overview of the latent space and generative flow is given in Figure 4, and the complete pipeline is illustrated in Figure 10.

### 2.1 Dataset and Clinical Context

The experimental data were obtained from the MIT-BIH Arrhythmia Database, a widely adopted benchmark for cardiac arrhythmia research comprising 48 half-hour two-channel ambulatory ECG recordings sampled at 360 Hz.^9^ Recordings were collected at Beth Israel Hospital from patients referred for extended ambulatory monitoring and represent a broad clinical spectrum of cardiac rhythms. Records 102, 104, 107, and 217 – which contain exclusively pacemaker-stimulated beats – were excluded, as paced QRS morphologies reflect artificial depolarization sequences and are not representative of spontaneous arrhythmia.

Beat annotations were mapped to the four standard clinical categories defined by the Association for the Advancement of Medical Instrumentation (AAMI),^33^ each corresponding to a distinct electrophysiological mechanism:

- **Class** 𝒩 **(Normal):** Sinus rhythm beats and bundle-branch block beats whose QRS morphology is characteristic for that patient. Depolarization originates at the sinoatrial node, propagates normally through the atrioventricular (AV) node and His-Purkinje system, and produces a narrow QRS complex with a regular PR interval.
- **Class** 𝒮 **(Supraventricular ectopic):** Premature beats originating above the bundle of His, encompassing premature atrial contractions (PACs) and junctional beats. 𝒮-class beats typically produce a narrow or slightly aberrant QRS complex preceded by an early, absent, or retrograde P wave. Clinically, frequent supraventricular ectopy may serve as an early marker for paroxysmal atrial fibrillation risk,^34^ and warrants monitoring in symptomatic patients.
- **Class** 𝒱 **(Ventricular ectopic):** Premature beats arising from an ectopic focus within the ventricular myocardium, bypassing the normal conduction system entirely. These produce a wide (*>* 120 ms), bizarrely shaped QRS complex with a compensatory pause and a discordant T wave. Frequent or complex ventricular ectopy – particularly multiform beats, bigeminy, or runs of non-sustained ventricular tachycardia – carries independent prognostic significance^13^ and may require clinical intervention, including antiarrhythmic therapy or catheter ablation.
- **Class** ℱ **(Fusion):** Beats resulting from near-simultaneous ventricular activation by both a normal sinus impulse and a ventricular ectopic impulse. The QRS morphology is intermediate between a normal and a ventricular beat, reflecting the hybrid electrical origin. Fusion beats indicate the presence of an active ventricular ectopic focus and are a key distinguishing feature in the differential diagnosis of wide-complex tachycardias.

Non-beat annotations and the unknown class were discarded. The resulting dataset exhibits pronounced class imbalance: Class 𝒩 constitutes 89.4% of all annotated beats, while the minority classes 𝒮 (2.7%), 𝒱 (7.2%), and ℱ (0.76%) together account for only 10.6% of the total (Figure 1). This distribution reflects the clinical reality of ambulatory monitoring, in which pathological events are inherently rare relative to a background of normal sinus rhythm, and constitutes the primary motivation for the generative augmentation pipeline developed in this work.

**Figure 1:**
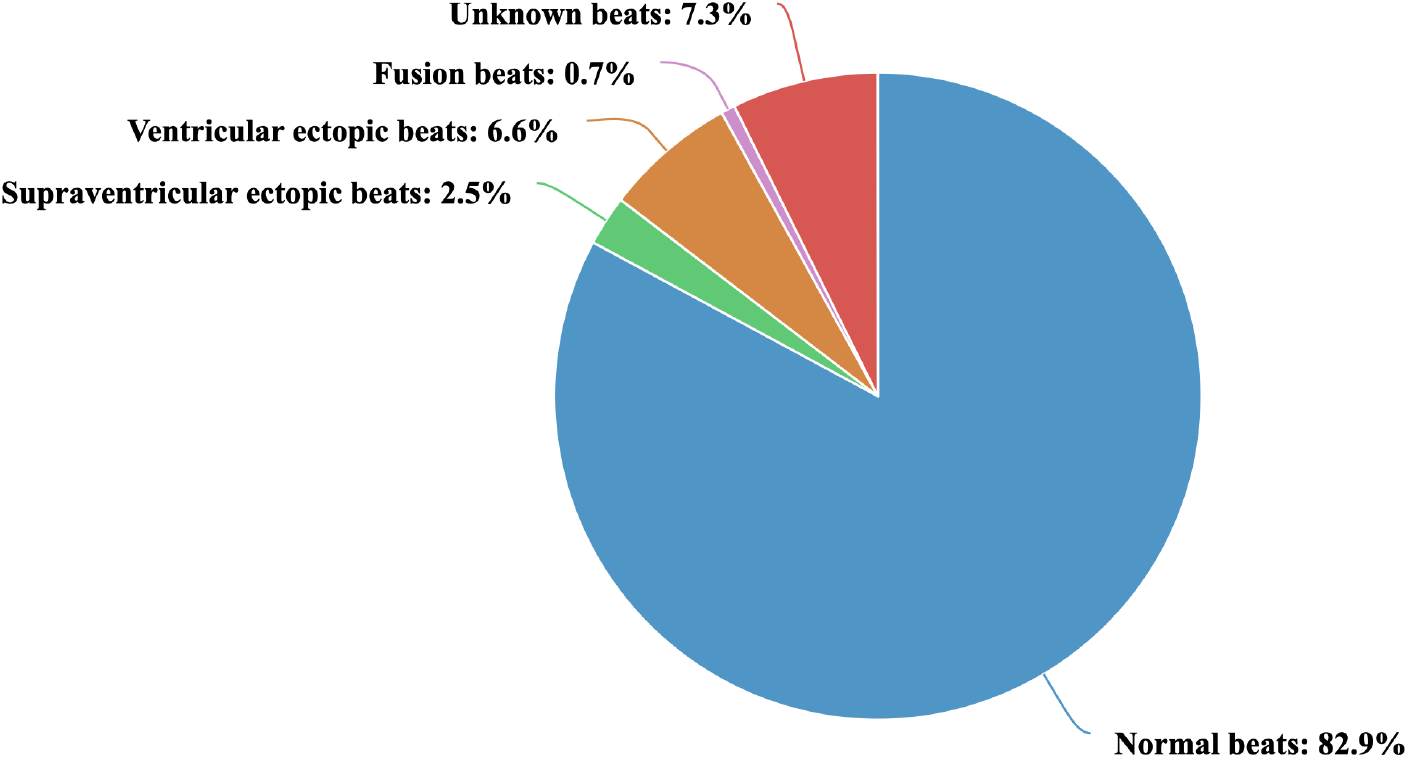
Distribution of beat annotations in the MIT-BIH Arrhythmia Database prior to filtering. Normal beats account for 82.9% of all raw annotations. The unknown class (7.3%) was subsequently discarded. After removing unknown and non-beat annotations, the four retained AAMI classes exhibit pronounced imbalance: Class 𝒩 constitutes 89.4% of retained beats, while the minority classes (𝒮, 𝒱, ℱ) together account for only 10.6%. This severe imbalance is the primary motivation for the generative augmentation pipeline.

**Figure 2:**
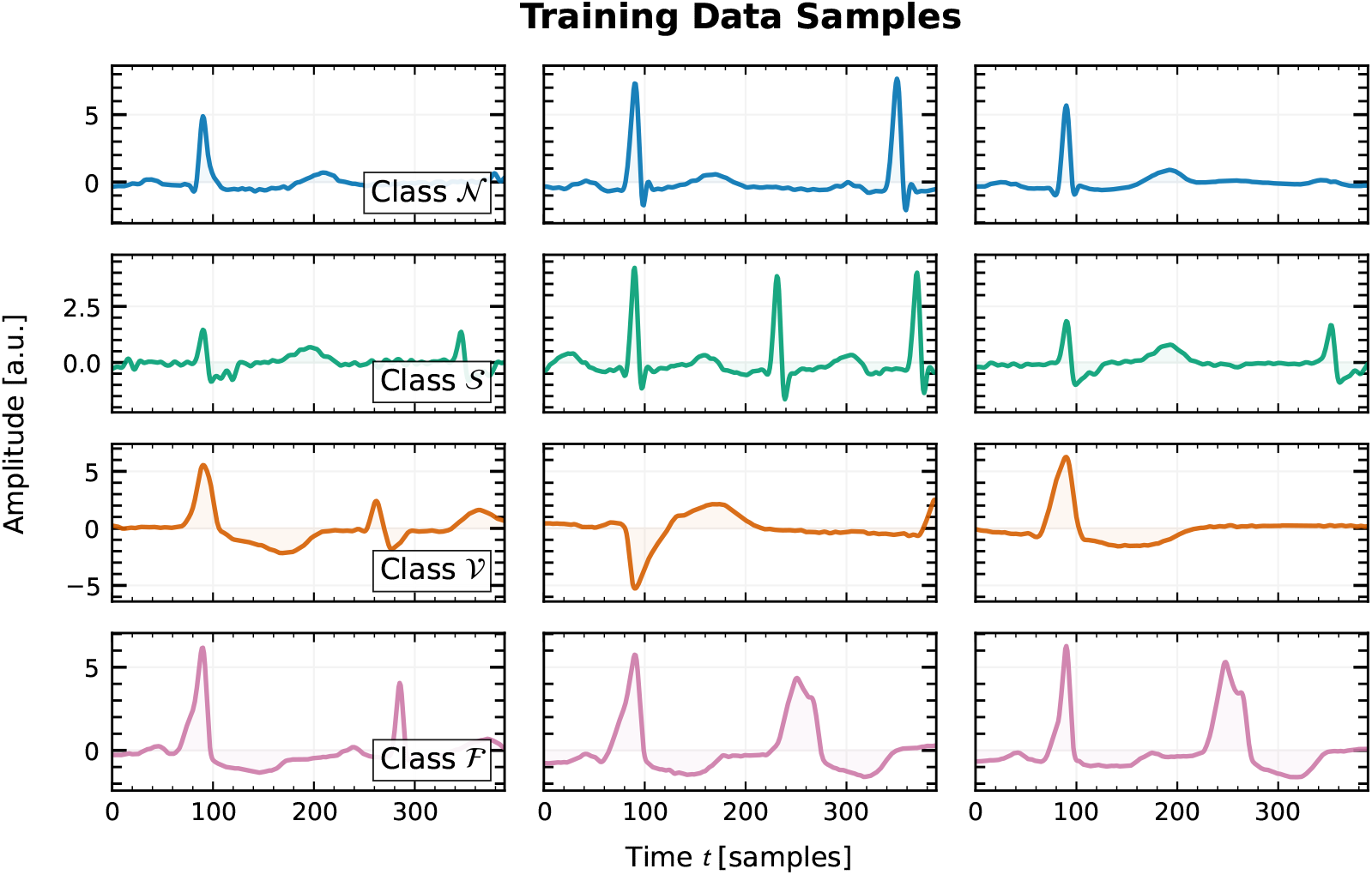
Representative beat segments drawn from the training split for each of the four AAMI classes. Each row shows three randomly selected samples. The vertical axis is amplitude in arbitrary units after global normalisation, and the horizontal axis is time in samples at 360 Hz. Class 𝒩 beats display a consistent narrow QRS morphology. Class 𝒮 beats are morphologically similar to Normal but exhibit subtle P-wave anomalies and slight QRS widening. Class 𝒱 beats are characterised by a wide, bizarrely shaped QRS complex and discordant T wave. Class ℱ beats show an intermediate morphology between Normal and Ventricular, reflecting their hybrid electrical origin.

The experimental design emulates a personalized wearable monitoring scenario in which a cardiac monitor is calibrated to a specific patient’s baseline rhythm before being deployed to detect arrhythmic events. An intra-patient temporal split is employed. For each record, the first 80% of beats are allocated to training and validation (80% training, 10% validation), and the final 10% to testing. This ensures that the classifier is evaluated on later segments of the same patient’s recording, which may exhibit morphological drift relative to the calibration window and thus constitutes a realistic test of personalized monitoring performance. Crucially, the split is applied independently within each record before any cross-record aggregation, so no beat from any patient’s test window is ever seen during training or validation of the generative models or the classifier. This design eliminates the intra-patient data leakage that arises when beats from the same recording are randomly shuffled and partitioned globally. After aggregation across all 44 retained records, the resulting real-beat counts per class and partition are given in Table 1. Figure 3 illustrates how the four augmentation ratios *ρ* ∈ {0.25, 0.50, 0.75, 1.00} translate into concrete training set sizes; at *ρ* = 1.00, for example, the pipeline synthesizes nearly 70,000 additional supraventricular samples from only 2,186 real 𝒮-class beats, underscoring the severity of the minority-class deficit.

**Table 1:** Real-beat counts per AAMI class and data partition after intra-patient temporal splitting across all 44 retained MIT-BIH records (80 / 10 / 10 split).

| Class | Train | Validation | Test |
| --- | --- | --- | --- |
| $\mathcal{N}$ (Normal) | 72,136 | 9,046 | 8,943 |
| $\mathcal{S}$ (Supraventricular) | 2,186 | 252 | 343 |
| $\mathcal{V}$ (Ventricular) | 5,527 | 718 | 764 |
| $\mathcal{F}$ (Fusion) | 708 | 53 | 42 |
| **Total** | **80,557** | **10,069** | **10,092** |

**Figure 3:**
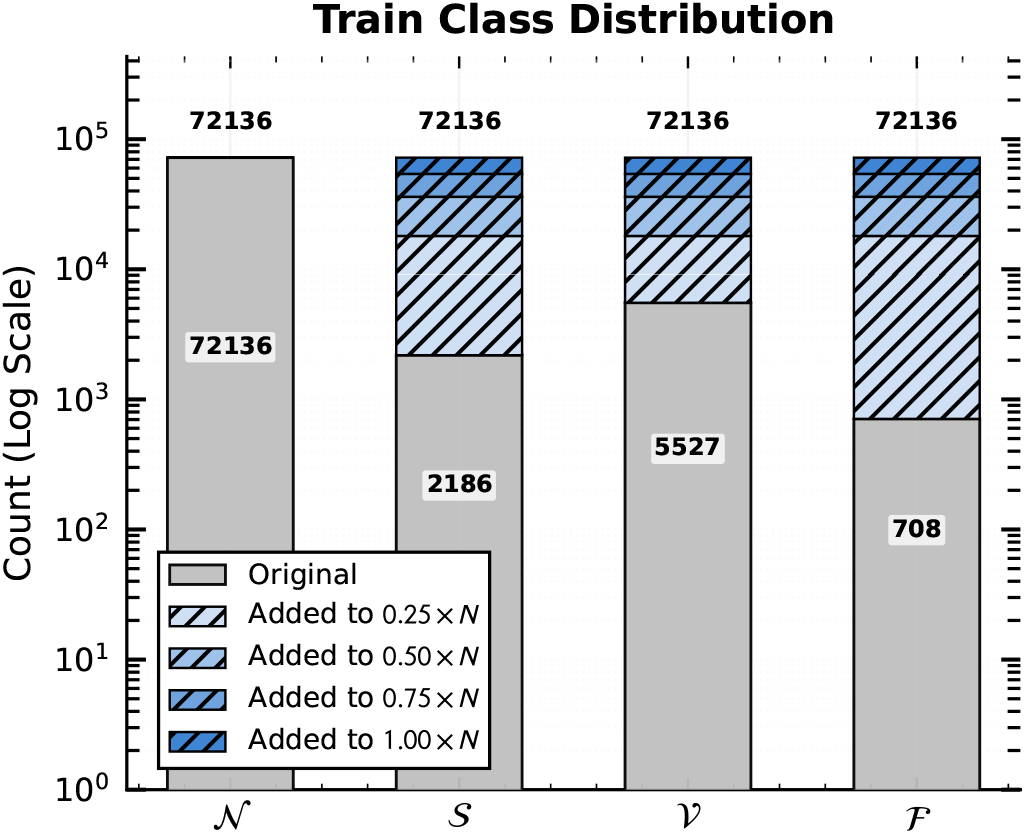
Training set class counts before and after synthetic augmentation, shown on a logarithmic scale. Grey bars represent real beats; blue hatched layers show cumulative synthetic beats added at each augmentation ratio *ρ* ∈ *{*0.25, 0.50, 0.75, 1.00*}*, where *ρ* = 1.00 brings all minority classes to parity with the *N* = 72,136 majority-class count. Class ℱ has fewer than 1% as many real beats as Class 𝒩, making it the most dependent on synthetic data at every ratio.

**Figure 4:**
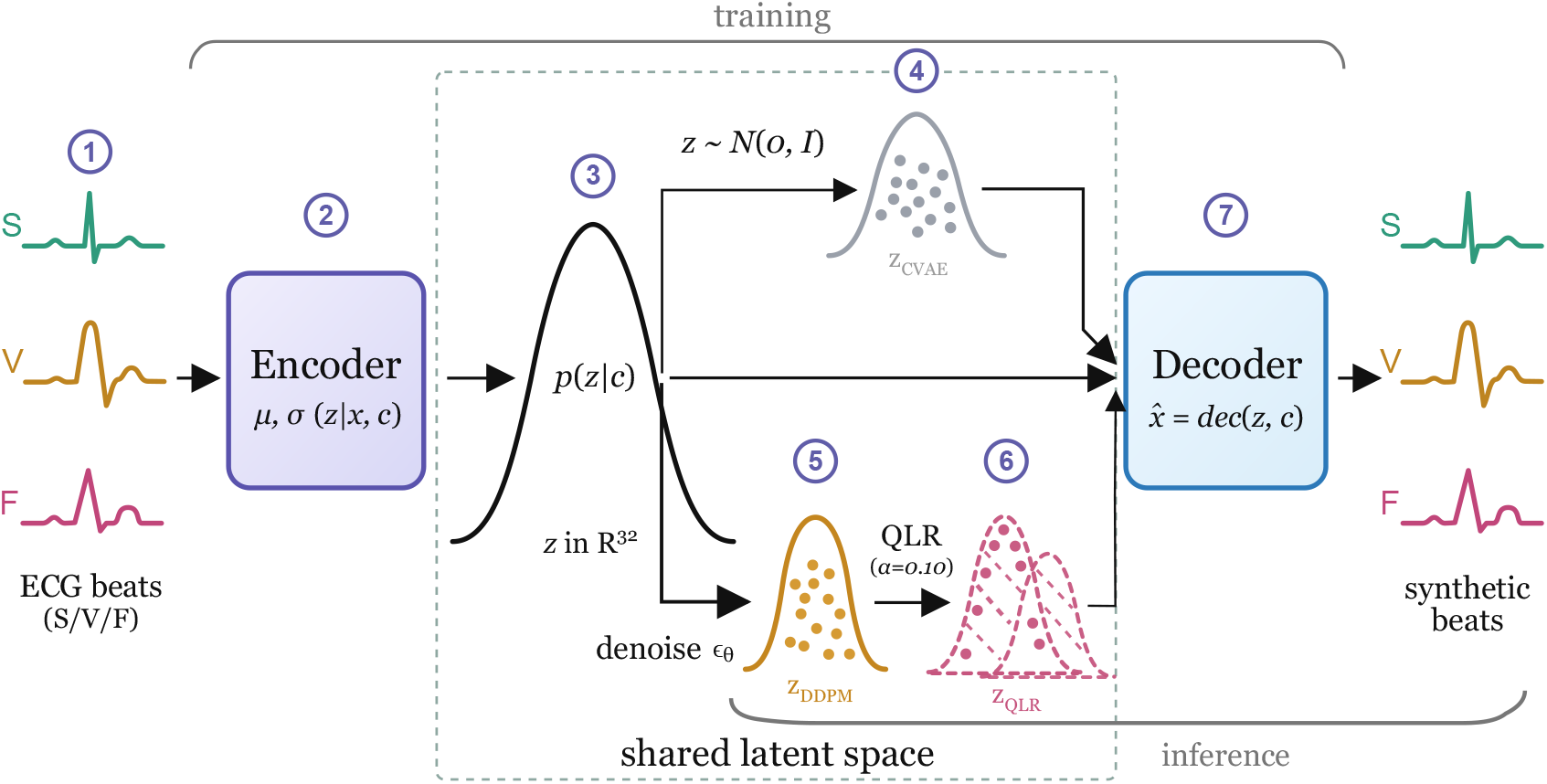
Overview of the cVAE latent space and its role in the generative pipeline. The encoder maps input ECG beats (𝒮, 𝒱, ℱ) into the shared 32-dimensional latent space via the class-conditional posterior *µ, σ*(*z x, c*) (steps 1–3). For cVAE-only generation (step 4), latents are sampled unconditionally from 𝒩 (0, *I*) with class conditioning applied at the decoder only. At inference, the DDPM denoiser *ϵ*_*θ*_ generates class-conditional latent samples (step 5), the QLR module applies a bounded distributional correction (step 6), and the shared decoder reconstructs synthetic ECG beats (step 7).

**Figure 5:**
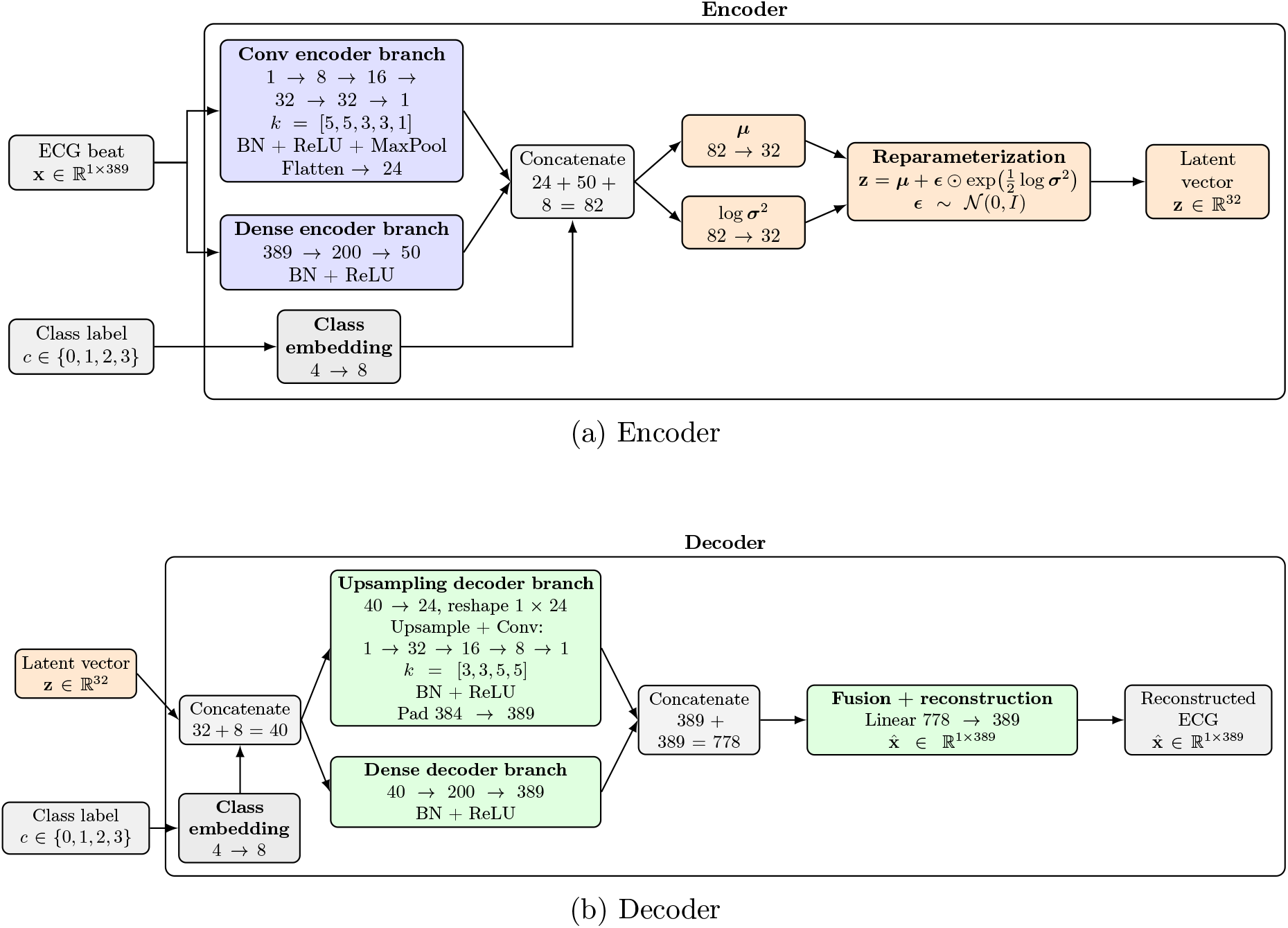
Hybrid conditional variational autoencoder architecture. (a) Encoder: the input ECG beat is processed through a convolutional branch for local morphological feature extraction and a dense branch for global waveform representation, while a learned class embedding provides conditional information. The merged representation is projected into the latent mean *µ* and log-variance log *σ*^2^, from which the latent code is sampled via the reparameterization trick. (b) Decoder: the latent code and class embedding are concatenated and decoded through an upsampling-convolution branch and a dense reconstruction branch. Their outputs are fused to reconstruct the ECG beat.

**Figure 6:**
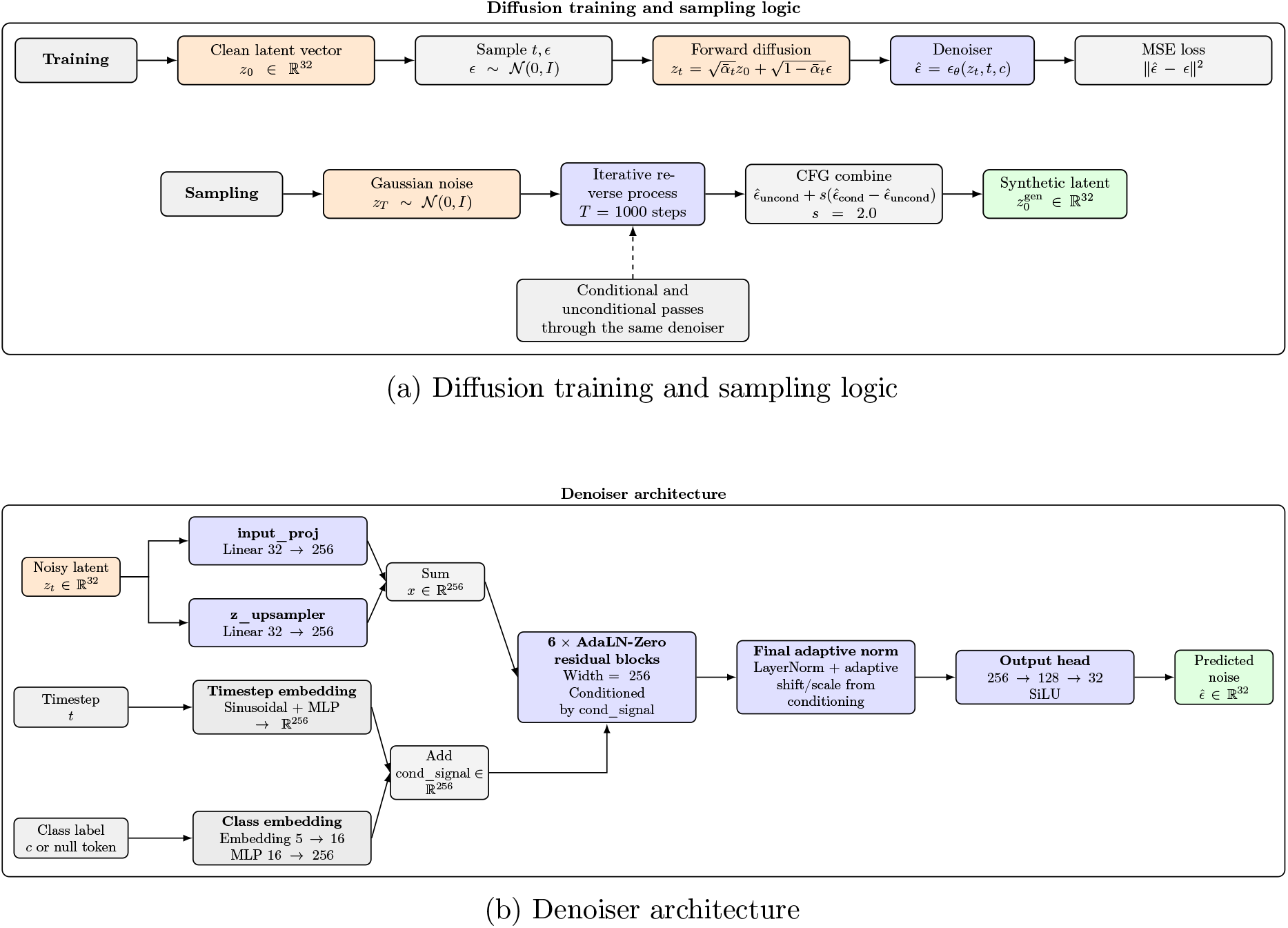
Class-conditional latent DDPM used to model minority-class cVAE latents. (a) During training, Gaussian noise is added to *z*_0_ at timestep *t* to form *z*_*t*_, and the denoiser *ϵ*_*θ*_ is optimized with the noise-prediction loss 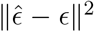. During sampling, reverse diffusion with classifier-free guidance (*s* = 2.0) recovers a synthetic latent 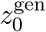. (b) The noisy latent *z*_*t*_ ∈ ℝ^32^ is projected through `input_proj` and `z_upsampler` and summed into a 256-dimensional hidden representation. Sinusoidal timestep and class embeddings are added to form the conditioning vector used by six AdaLN-Zero residual blocks. A final adaptive normalization and output head (256 →128 →32) produce the predicted noise 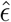.

### 2.2 Signal Preprocessing

A second-order Butterworth bandpass filter with cutoff frequencies of 0.5 Hz and 45 Hz was applied to each ECG channel before any further processing. The lower cutoff eliminates baseline wander – the low-frequency drift artifact produced by patient respiration and electrode movement that is especially prevalent in ambulatory recordings – while the upper cutoff suppresses high-frequency skeletal muscle (EMG) noise. These bounds are consistent with standard clinical ECG processing guidelines and preserve all diagnostically relevant waveform components: the P wave (0.5–4Hz), the QRS complex (3–40Hz), and the T wave (0.5–10Hz), including the high-frequency notches within the QRS that may signal bundle-branch conduction abnormalities.

Individual beats were extracted using annotation-guided windowing, centered on each annotated R-peak with a fixed window of 389 samples (≈0.25 s pre-peak, 0.83 s post-peak).^35^ The window boundaries were chosen to guarantee complete capture of the P-QRS-T complex across all four AAMI beat classes. The extended post-R-peak interval of 0.83 s accommodates prolonged QT intervals – as seen in patients receiving QT-prolonging antiarrhythmic agents or in congenital long QT syndrome – ensuring that T-wave offset is preserved in every extracted beat.

Global Z-score normalization was applied using the mean and standard deviation computed from the training set (*µ*_train_, *σ*_train_), rather than on a per-beat basis. Local per-beat normalization would destroy relative amplitude information that is clinically diagnostic. Ventricular ectopic beats are frequently identified by high-voltage, wide QRS complexes, and ischemic changes present as subtle but critical ST-segment depressions or elevations. By normalizing with training-set statistics, inter-class amplitude relationships are preserved, maintaining the voltage features that differentiate arrhythmia types and are essential for any deep learning model trained on this representation.

### 2.3 Hybrid Conditional Variational Autoencoder (cVAE)

A variational autoencoder (VAE)^36^ learns a compressed representation of its inputs: the encoder maps each input to a point in a low-dimensional latent space, and the decoder reconstructs the original signal from that point. Building upon prior VAE architectures for ECG representation learning,^37^ the conditional variant (cVAE) developed here additionally conditions both encoder and decoder on a class label, organizing the latent space by arrhythmia type. Performing all subsequent generation in this latent space – rather than directly on the raw 389-sample signal – allows the downstream diffusion model to operate in a far lower-dimensional and more structured domain, making both training and sampling substantially more efficient than raw-signal generation. The first stage of the pipeline therefore compresses each 389-sample ECG beat into a compact 32-dimensional latent vector *z* using a spectral-guided cVAE.

The encoder adopts a hybrid architecture: a convolutional branch extracts local morphological structure through four 1D feature-extraction blocks (each with batch normalization, ReLU activation, and max pooling), followed by a 1 *×* 1 projection convolution; in parallel, a dense branch processes the full beat through two fully connected layers. A learned class embedding of dimension 8 is concatenated with the outputs of both branches, and the merged representation is used to predict the latent mean *µ* and log-variance log *σ*^2^. The decoder follows a complementary hybrid structure with upsampling-convolutional and dense reconstruction paths, fused through a final linear layer.

The cVAE is trained with an ELBO extended with a spectral consistency term:

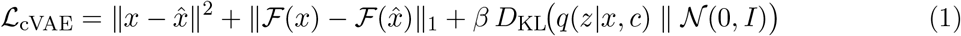

where ℱ (·) denotes the discrete Fourier transform magnitude. The spectral loss term explicitly penalizes frequency-domain discrepancies between real and reconstructed beats – particularly important for ECG signals, where diagnostic information spans heterogeneous frequency bands. The high-frequency notches of the QRS complex must be reproduced faithfully alongside the low-frequency arc of the T wave. Without this term, reconstruction tends to over-smooth high-frequency morphological detail in favor of minimizing the global MSE.

The KL weight *β* is linearly annealed from 0 to 0.05 over the first 40 training epochs and then held constant, preventing posterior collapse^38^ (the failure mode in which the encoder ignores the input and maps all observations to the prior). The model is trained for 130 epochs using the Adam optimizer^39^ (learning rate 10^−3^, gradient clipping with maximum norm 1.0). At inference, the encoder mean *µ*(*x, c*) is used in place of sampled posterior latents, yielding a deterministic and reproducible latent representation for the downstream diffusion model.

### 2.4 Class-Conditional Latent Diffusion Model (DDPM)

A diffusion model learns to reverse a gradual noising process: during training, Gaussian noise is progressively added to a clean latent vector *z*_0_ over *T* timesteps until it approximates an isotropic Gaussian *z*_*T*_ ∼ 𝒩 (0, *I*). The model learns to predict the added noise at each step, so that at inference it can begin from a random Gaussian sample and iteratively denoise it back into a realistic latent vector.^18,19^ Conditioning on a class label steers this process toward the latent distribution of the target arrhythmia class, enabling targeted minority-class synthesis.

The second stage trains a class-conditional latent DDPM on the cVAE latent representations to model and sample from the class-specific latent distributions of minority arrhythmia beats. The denoiser is a residual MLP with AdaLayerNorm-Zero (AdaLN-Zero) conditioning:^40^ a learned class embedding and a sinusoidal timestep embedding are projected into a shared conditioning signal that modulates six residual blocks through scale and shift parameters. The latent input *z* is first projected into the model dimension R^256^ through two parallel linear layers with independent initializations, whose outputs are summed to form the hidden representation *x*, after which the representation is processed by six AdaLN-Zero residual blocks.

Training latents are standardized dimension-wise using training-set statistics (*µ*_*z*_, *σ*_*z*_) before being passed to the model. The forward diffusion process corrupts the standardized latents over *T* = 1000 steps under a linear variance schedule, *β*_*t*_ ∈ [ 10^−4^, 10^−2^ ], uniformly spaced. The model is optimized under the standard noise-prediction objective:

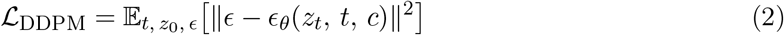

where *ϵ* ∼ 𝒩 (0, *I*) is the noise target and *ϵ*_*θ*_ is the learned denoiser.

Classifier-free guidance (CFG)^41^ is applied by randomly replacing the class conditioning with a null token with probability 0.1 during training, and using a guidance scale of 2.0 at inference. A class-balanced `WeightedRandomSampler` ensures equal minority-class exposure during training. The model is optimized with Adam^39^ (learning rate 10^−4^, gradient clipping with maximum norm 1.0) and an exponential moving average (EMA, decay 0.9999) of the model parameters. All reported samples are drawn from the EMA-smoothed model.

### 2.5 Quantum Latent Refinement (QLR)

The QLR module operates as a post-processing refinement step: after the DDPM generates a synthetic latent that approximates the target class distribution, the QLR applies a bounded correction to shift it toward higher-density regions of the real latent manifold – guided by the displacement from real examples – without altering it so drastically that the underlying beat morphology is lost. This refinement is implemented using a parameterized quantum circuit (PQC), whose entanglement structure introduces inter-dimensional correlations that are structurally distinct from those of a classical network of equivalent parameter count.

Even with classifier-free guidance, DDPM-generated minority-class latents may still occupy lower-density regions of the real latent manifold. This is particularly pronounced for morphologically complex classes such as ventricular ectopy, whose latent distribution is broad and multimodal. To reduce this mismatch, a QLR module is trained separately for each minority class (𝒮, 𝒱, and ℱ) using an 8-qubit PQC.^21^

Prior to circuit embedding, each 32-dimensional latent is mapped to the quantum domain [−1, 1]^32^ via an affine transformation *ϕ*(·) defined by the 1st and 99th percentiles of the real class-specific latent bank. This per-class calibration ensures that the data-embedding rotations operate within the valid angular range of the quantum gates. The circuit architecture consists of three components:

1. **Single data-embedding stage**. The 32 normalized latent dimensions are embedded once at the circuit input using two *R*_*Y*_ and two *R*_*Z*_ rotations per qubit. Data re-uploading at intermediate layers^42^ is not employed, keeping the circuit shallow to mitigate barren plateau phenomena.^22^
2. **Trainable rotation–entanglement stack**. Six layers follow, each applying a general Rot gate (*R*_*z*_-*R*_*y*_-*R*_*z*_) to every qubit, followed by a brick-pattern CNOT entanglement scheme: even layers connect qubit pairs (0, 1), (2, 3), (4, 5), (6, 7); odd layers connect (1, 2), (3, 4), (5, 6) with a wrap-around (7, 0) connection. Every third layer adds stride-2 skip entanglement.
3. **Multi-observable readout**. The circuit returns 32 expectation values: Pauli-*Z*, Pauli-*X*, Pauli-*Y*, and nearest-neighbor *Z* ⊗ *Z* per qubit location.

The resulting circuit is illustrated in Figure 7.

**Figure 7:**
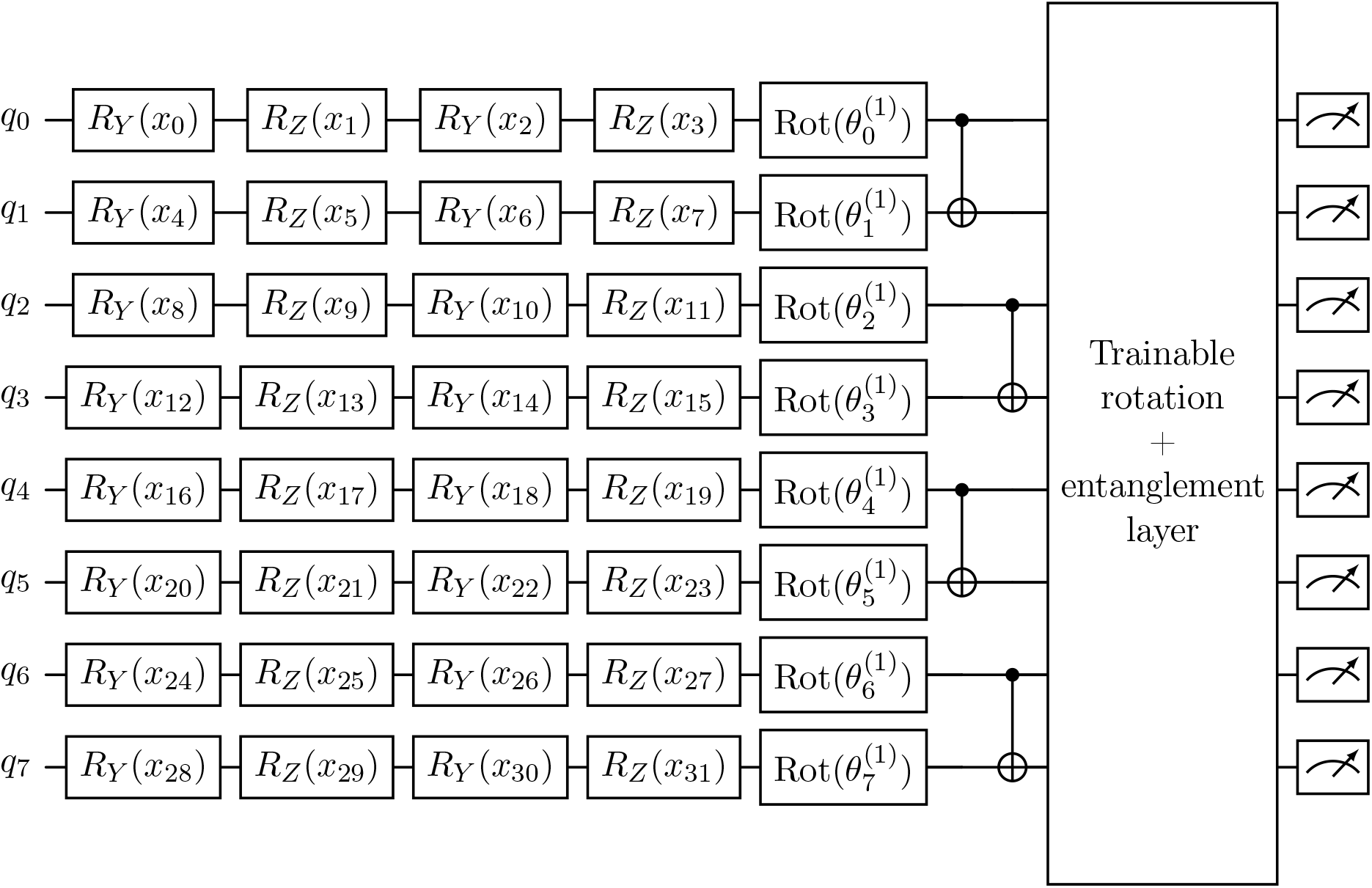
Parameterized quantum circuit used in the Quantum Latent Refinement (QLR) module. A normalized 32-dimensional latent vector is embedded once across 8 qubits using four angle encodings per qubit: *R*_*Y*_ (*x*_4*q*_), *R*_*Z*_(*x*_4*q*+1_), *R*_*Y*_ (*x*_4*q*+2_), and *R*_*Z*_(*x*_4*q*+3_). This is followed by a trainable rotation-entanglement layer repeated for 6 layers. In each layer, every qubit receives a general single-qubit Rot gate, followed by an alternating brick-pattern CNOT entanglement scheme; odd layers include wrap-around entanglement, and every third layer includes stride-2 skip entanglement. The circuit readout combines Pauli-*Z*, Pauli-*X*, Pauli-*Y*, and nearest-neighbor *Z* ⊗ *Z* expectation values to form the 32-dimensional refinement signal applied in latent space.

The circuit output is standardized per sample and applied as a bounded residual correction:

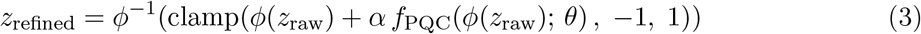

with correction scale *α* = 0.10. The small scale factor constrains the QLR to regularize the synthetic distribution rather than overwrite the morphological content encoded by the DDPM – preserving the beat structure while improving distributional alignment. The full module and its training objective are illustrated in Figure 8.

**Figure 8:**
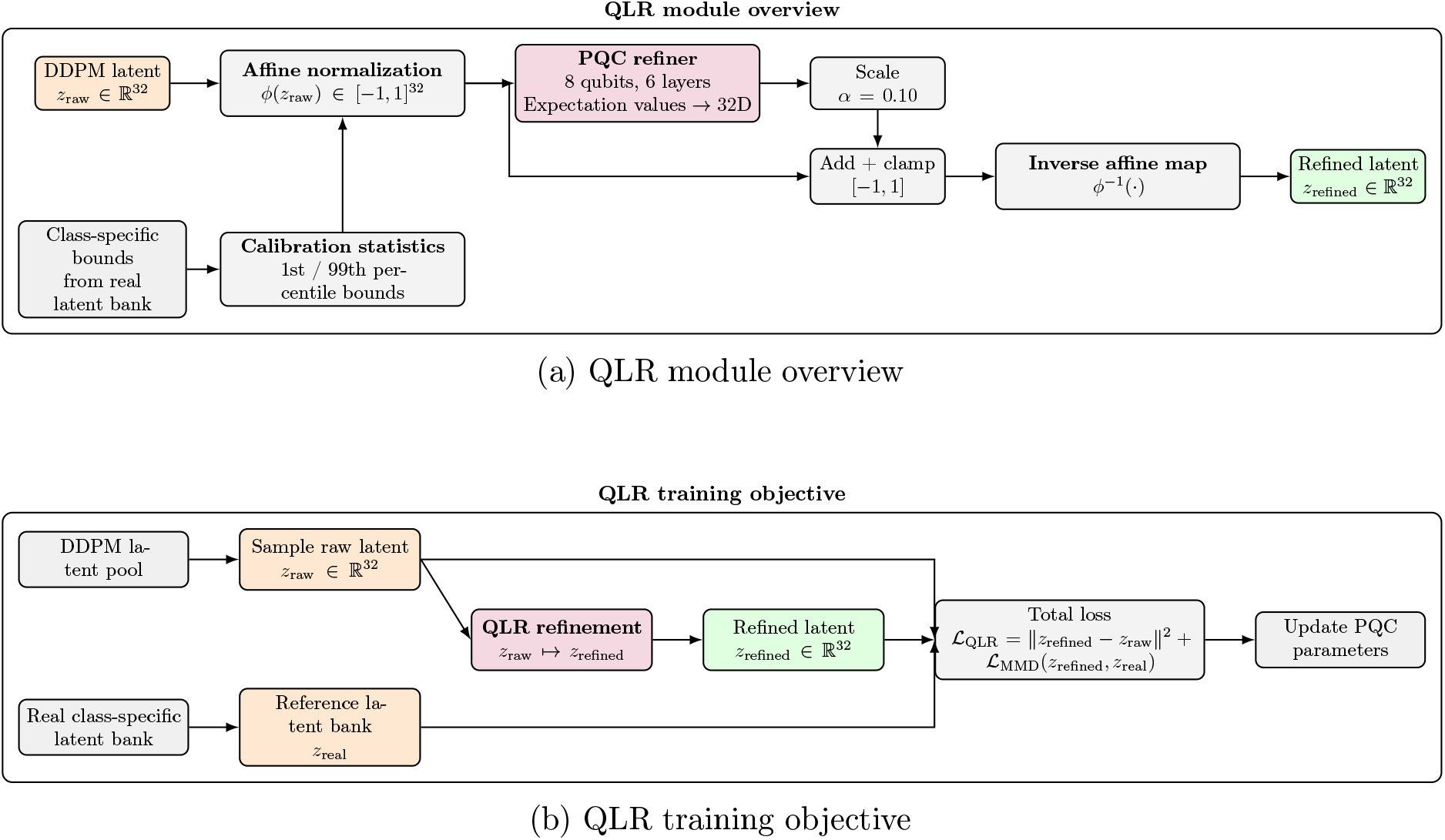
Quantum Latent Refinement (QLR) module used to refine DDPM-generated minority-class latents. (a) QLR module overview: the raw DDPM latent *z*_raw_ ∈ ℝ^32^ is mapped to the normalized quantum domain through an affine transformation *ϕ*(*z*_raw_) ∈ [ −1, 1]^32^ using class-specific bounds derived from the real latent bank. A parameterized quantum circuit (PQC) produces a refinement signal that is scaled by *α* = 0.10, added to the normalized latent, and clamped to the interval [ −1, 1]. The refined representation is then mapped back to the original latent space through the inverse transform *ϕ*^−1^ to obtain *z*_refined_. (b) QLR training objective: a raw latent sampled from the DDPM pool is refined by the QLR module and compared both to its original DDPM source through the stay loss ℒ_stay_ = ∥*z*_refined_ − *z*_raw_∥^2^ and to the real class-specific latent bank through the MMD loss ℒ_MMD_(*z*_refined_, *z*_real_). The total QLR objective is ℒ_QLR_ = ℒ_stay_ + ℒ_MMD_.

QLR training uses a two-term unsupervised objective:

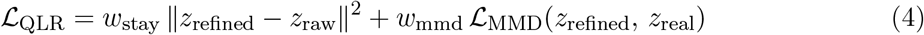

with *w*_stay_ = 1.0 and *w*_mmd_ = 1.0. The stay-loss penalizes large displacements from the DDPM origin, preserving the morphological characteristics encoded in each latent vector. The MMD term guides the refined distribution toward the real class-specific latent bank. It is computed as the squared Maximum Mean Discrepancy^43^ with an adaptive-bandwidth RBF kernel:

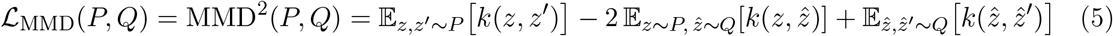

where *k*(*z, z*^*′*^) = exp −∥*z* − *z*^*′*^∥^2^*/*(2*σ*^2^) is the RBF kernel and *σ* is set to the median pairwise distance of the real latent bank (the median heuristic^43^). A value of *L*_MMD_ = 0 indicates that the two distributions are identical; larger values reflect systematic displacement of the synthetic cloud from the real manifold. In practice, the following finite-sample estimator is used:

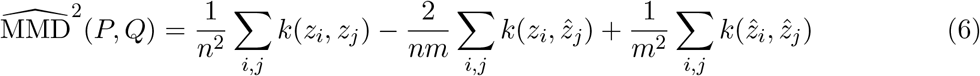

where *n* and *m* are the number of real and synthetic latent samples, respectively. Per-class training uses Adam^39^ at learning rate 2 *×* 10^−3^ with cosine annealing^44^ (*η*_min_ = 5 *×* 10^−4^), a pool of 2048 DDPM-generated samples refreshed every 5 epochs to prevent overfitting to a fixed synthetic set, early stopping with patience 20, and gradient clipping (norm 1.0).

The PQC is implemented in PennyLane^45^ using the `default.qubit` CPU-based simulator.

### 2.6 Downstream Classifier and Evaluation Protocol

The downstream classifier is a 1D adaptation of MobileNetV2,^32^ chosen for its inverted residual architecture and depth-wise separable convolutions, which together yield a compact parameter footprint suitable for resource-constrained wearable platforms where model storage and inference latency are critical. The network begins with a 1D convolutional stem followed by a sequence of inverted residual blocks. The classification head receives concatenated adaptive average-pooled and max-pooled features, enabling the model to integrate global rhythmic context with local morphological detail (Figure 9).

**Figure 9:**
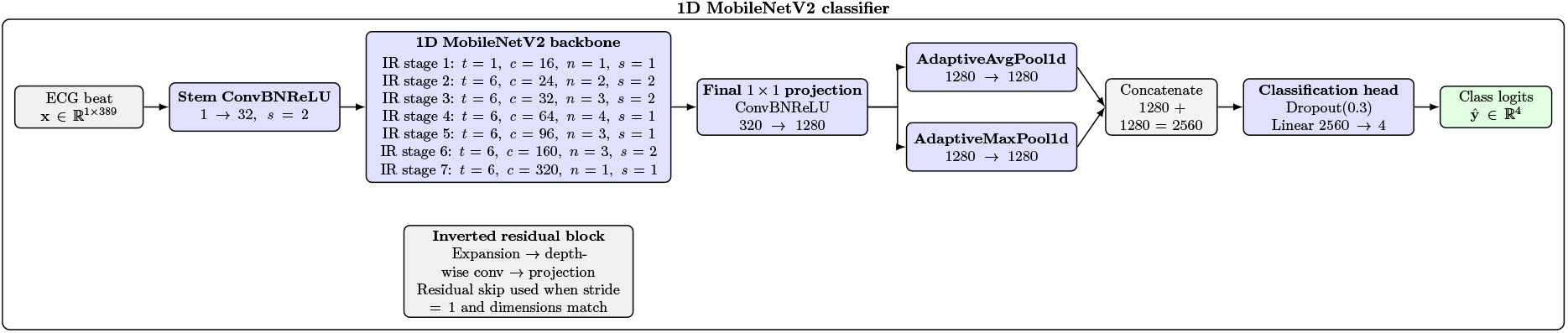
1D MobileNetV2 classifier used for downstream arrhythmia classification. An input ECG beat is first processed by a convolutional stem (1 →32, stride 2), followed by a stack of seven MobileNetV2 inverted residual stages parameterized by expansion ratio *t*, output channels *c*, number of blocks *n*, and stride *s*. A final 1 ×1 projection maps the backbone features from 320 to 1280 channels. Global feature aggregation uses both adaptive average pooling and adaptive max pooling; the resulting 1280-dimensional descriptors are concatenated into a 2560-dimensional representation. A dropout layer (*p* = 0.3) and final linear layer (2560 4) produce the class logits.

**Figure 10:**
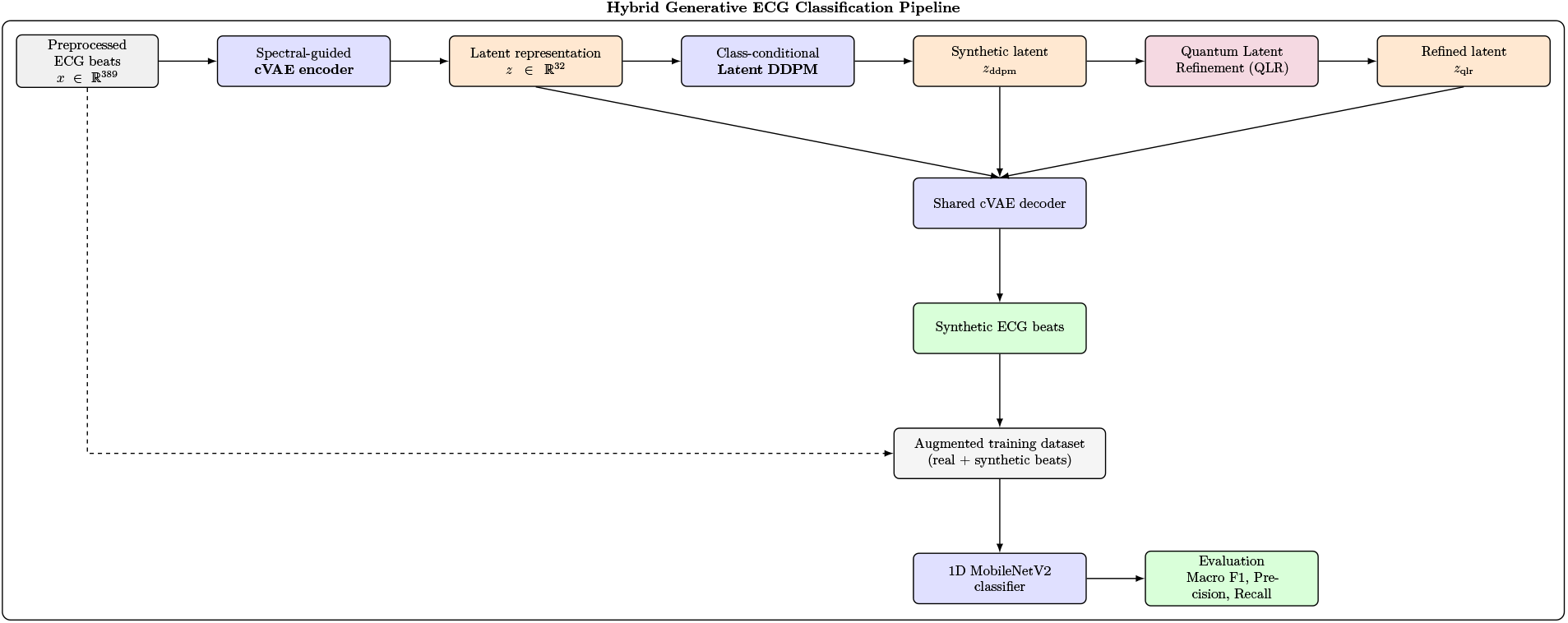
Overview of the proposed hybrid generative framework for imbalanced ECG classification. Preprocessed ECG beats are encoded into a 32-dimensional latent space using a spectral-guided conditional variational autoencoder (cVAE). The latent representation serves as the basis for direct cVAE generation, class-conditional latent diffusion (DDPM), and optional Quantum Latent Refinement (QLR). A shared cVAE decoder maps latent vectors back to the signal domain to produce synthetic ECG beats, which are combined with real beats to form an augmented training dataset. A lightweight 1D MobileNetV2 classifier is then trained on the augmented dataset and evaluated using Macro F1, precision, and recall.

Residual class imbalance in the augmented training set is addressed through inverse-frequency weighted cross-entropy (Eq. 7):

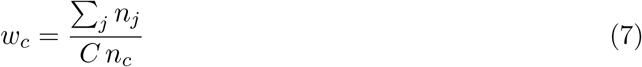

where *n*_*c*_ is the number of training samples in class *c* after augmentation and *C* is the number of classes. The weights are recomputed for each augmentation ratio *ρ*, so that the loss function reflects the augmented class distribution at every experimental condition. Weighted cross-entropy was preferred over focal loss to provide a transparent and interpretable imbalance correction. The interaction between explicit class up-weighting and synthetic augmentation is evaluated empirically rather than disentangled. The classifier is trained with Adam^39^ at learning rate 10^−3^, retaining the checkpoint with the best validation Macro F1, for a maximum of 100 epochs.

The full pipeline is evaluated across five independent random seeds and four augmentation ratios *ρ* ∈ *{*0.25, 0.50, 0.75, 1.00*}*, where *ρ* = 1.00 upsamples each minority class to match the majority Class 𝒩 count (72,136 beats). The five augmentation conditions are: (1) unaugmented baseline; (2) SMOTE; (3) cVAE-based sampling; (4) latent DDPM; and (5) DDPM+QLR. For cVAE generation, latent vectors are sampled from 𝒩 (0, *I*) with class conditioning applied at the decoder only, rather than through a learned class-conditional prior. This means the cVAE latent space is not explicitly structured by class, which limits the fidelity of unconditional latent samples relative to DDPM-generated ones.

A matched ablation compares the QLR against a classical MLP refiner (162 parameters) trained under identical conditions – same optimizer, learning-rate schedule, pool refresh policy, patience, and gradient clipping – on Class 𝒱. Statistical significance between methods is assessed via paired *t*-test (`scipy.stats.ttest_rel`) on the five seed-wise Macro F1 values. The entire pipeline is implemented in PyTorch,^46^ with GPU acceleration for the classical stages and CPU-based quantum simulation for the QLR module.

The primary evaluation metric is Macro F1 (Section 1). Per-class precision, recall, and confusion matrices are reported to characterize the clinical utility of each method, with particular attention to the ventricular class, where the sensitivity to specificity tradeoff carries direct prognostic implications.

## 3 Results

### 3.1 Synthetic Data Quality

#### 3.1.1 Training Convergence

Both generative models exhibited stable convergence (Figure 11). The cVAE training loss showed an expected initial rise during the KL annealing window (epochs 0–40) – as the posterior is progressively pushed toward the prior – followed by monotonic descent to a final training loss of approximately 1.67 (validation: 1.80). The modest and consistent train-validation gap confirms that the cVAE generalizes well without significant overfitting. The latent DDPM converged over 200 epochs from an initial noise-prediction loss near 0.42 to a final value of approximately 0.16 (validation: 0.19). The EMA-smoothed model provided a stable and smooth optimization trajectory throughout.

**Figure 11:**
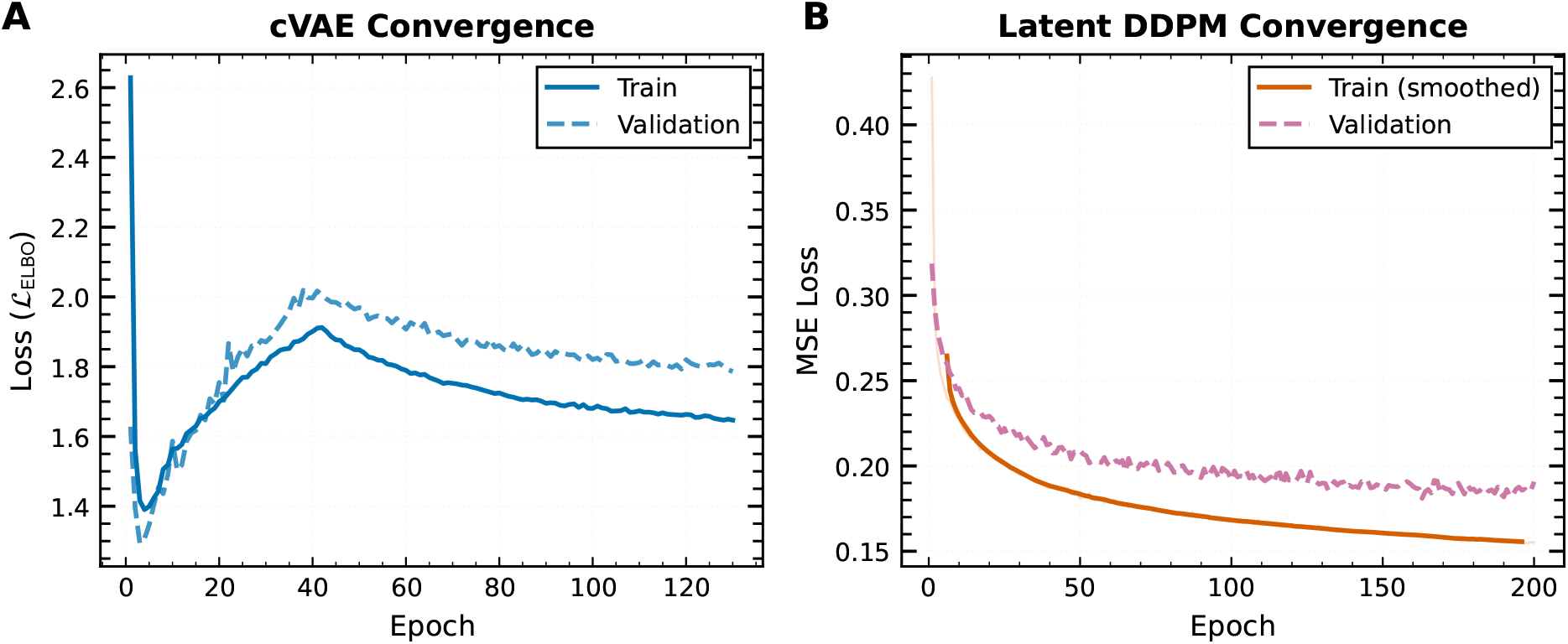
Training and validation convergence curves for the cVAE (Panel A) and latent DDPM (Panel B). The KL annealing-induced rise in the ELBO loss (Panel A, epochs 0–40) is characteristic of *β*-VAE training; the loss descends monotonically after annealing completes. The EMA-smoothed DDPM loss (Panel B) decreases consistently from ∼0.42 to ∼0.16 over 200 epochs.

#### 3.1.2 Morphological Fidelity of Synthetic Beats

Figures 12–14 compare the class-wise mean waveforms and one-standard-deviation envelopes of real versus synthetic beats for all three generative methods. Across all minority classes, synthetic mean waveforms closely track the real class means, confirming that the pipeline faithfully reproduces the dominant morphological signature of each arrhythmia type. The 𝒮-class synthetic beats reproduce the narrow QRS with subtle P-wave variation, the 𝒱-class beats capture the wide bizarrely-shaped QRS with biphasic deflection, and the ℱ-class beats preserve the hybrid morphology between Normal and Ventricular impulses. An exception worth noting is the DDPM cosine similarity for Class 𝒱 (0.9464), which is meaningfully lower than all other entries in Table 2. This reflects the high intra-class morphological variability of Ventricular ectopic beats: with a broad, multi-modal latent distribution, the class-mean waveform is a poor representative of individual beats, making cosine similarity of the mean a less informative fidelity measure for this class than root mean square deviation (RMSD).

**Figure 12:**
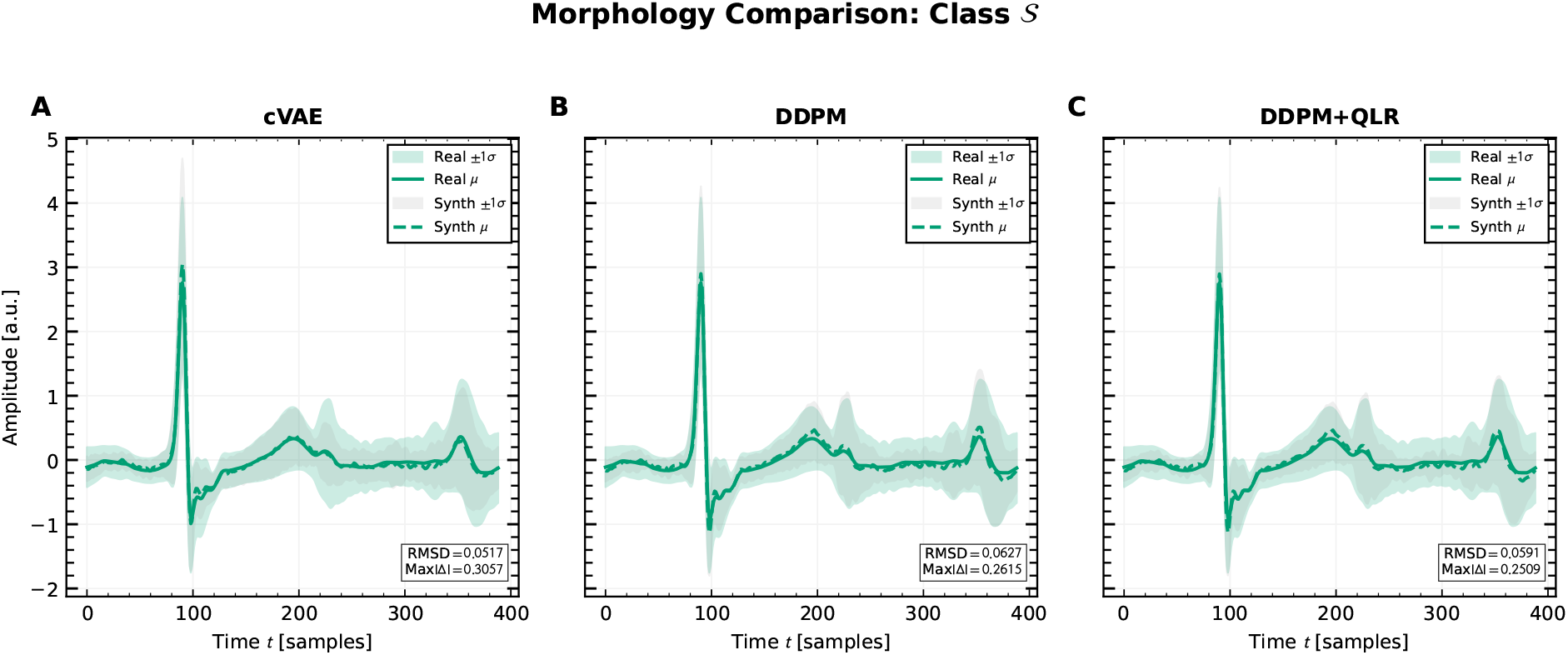
Morphology comparison for Class 𝒮. Real mean ±1*σ* envelope overlaid with synthetic statistics for cVAE (A), DDPM (B), and DDPM+QLR (C). The cVAE achieves the lowest RMSD (0.052) and highest cosine similarity (0.994). DDPM+QLR produces the smallest maximum absolute deviation (0.251).

**Figure 13:**
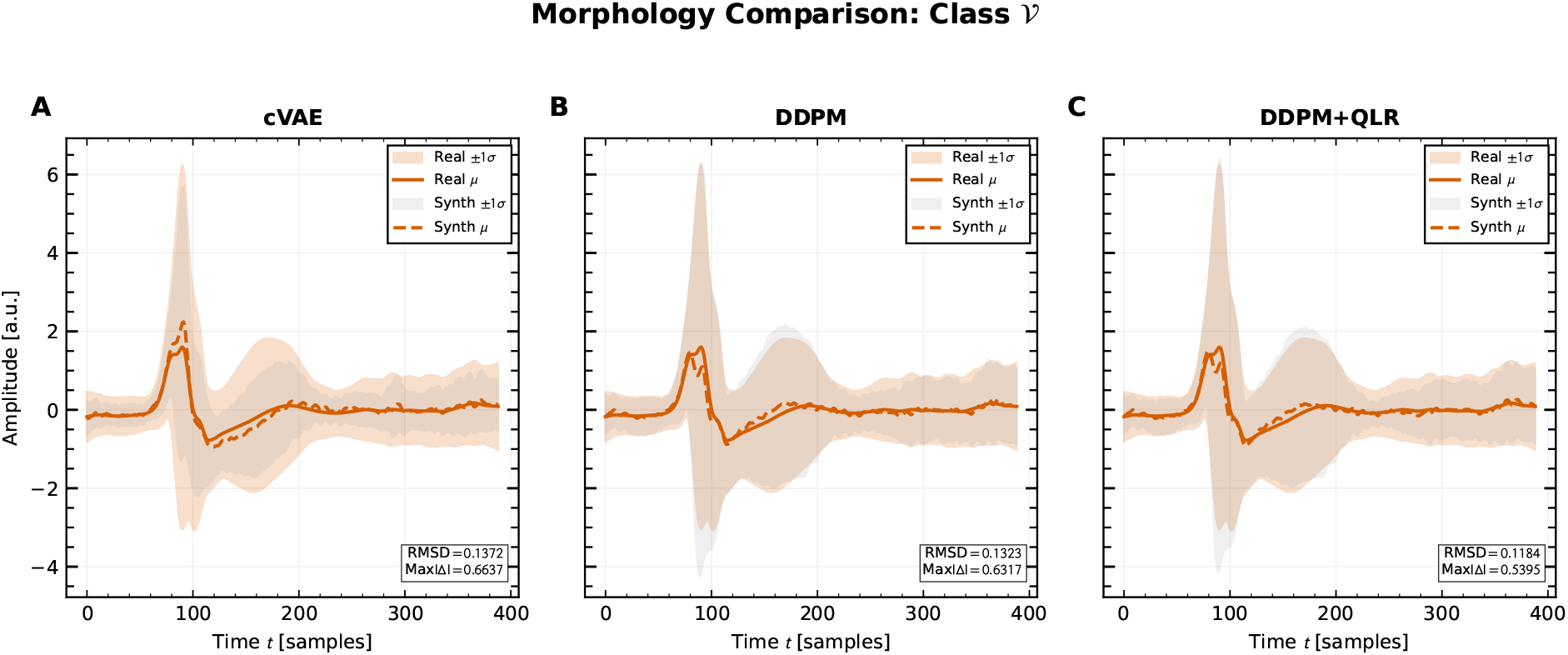
Morphology comparison for Class 𝒱. The Ventricular class exhibits the broadest ±1*σ* envelopes, reflecting its high morphological variability and multi-modal latent structure. DDPM+QLR achieves the best RMSD (0.118) and lowest maximum absolute deviation (0.540), indicating that quantum refinement provides its most consistent waveform-level contribution in this clinically critical class.

**Figure 14:**
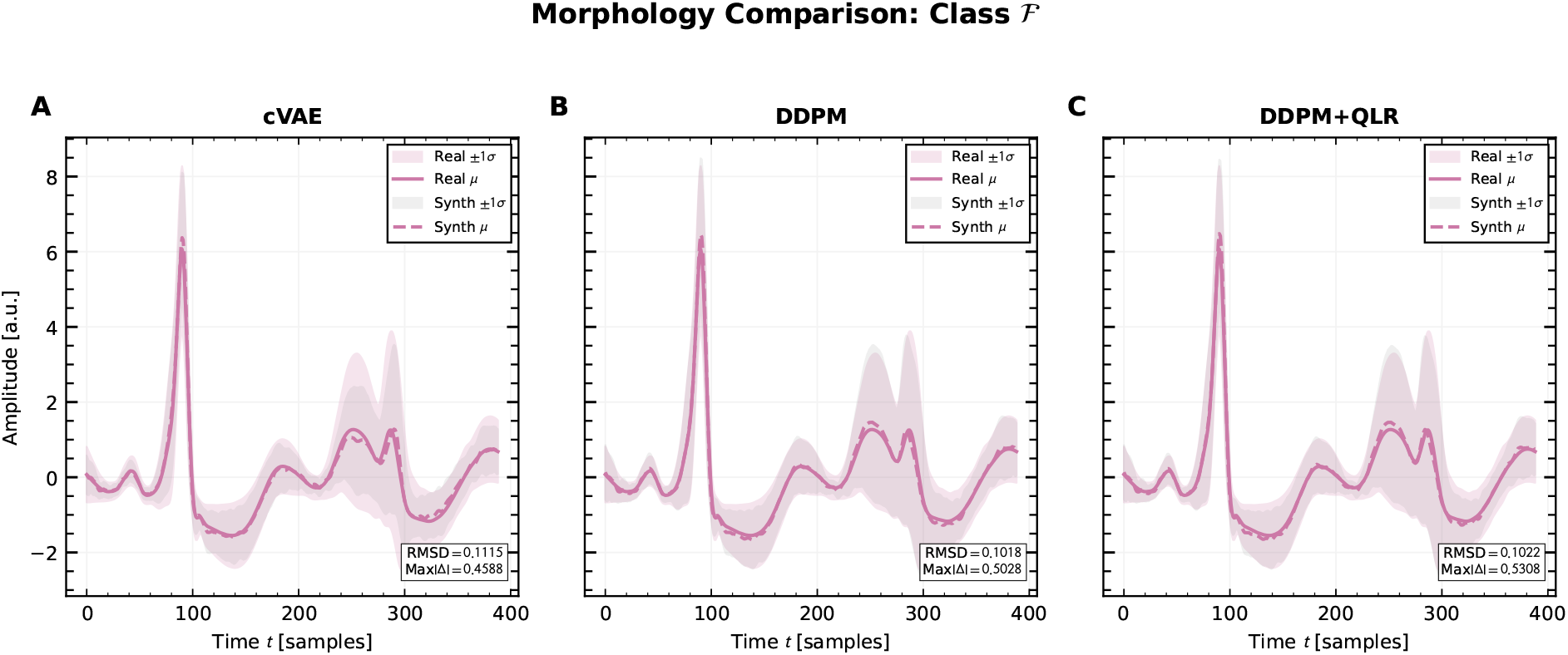
Morphology comparison for Class ℱ. All three methods capture the hybrid QRS morphology with similar fidelity. DDPM achieves the best RMSD (0.102) and cosine similarity (0.998) for this class.

**Table 2:**
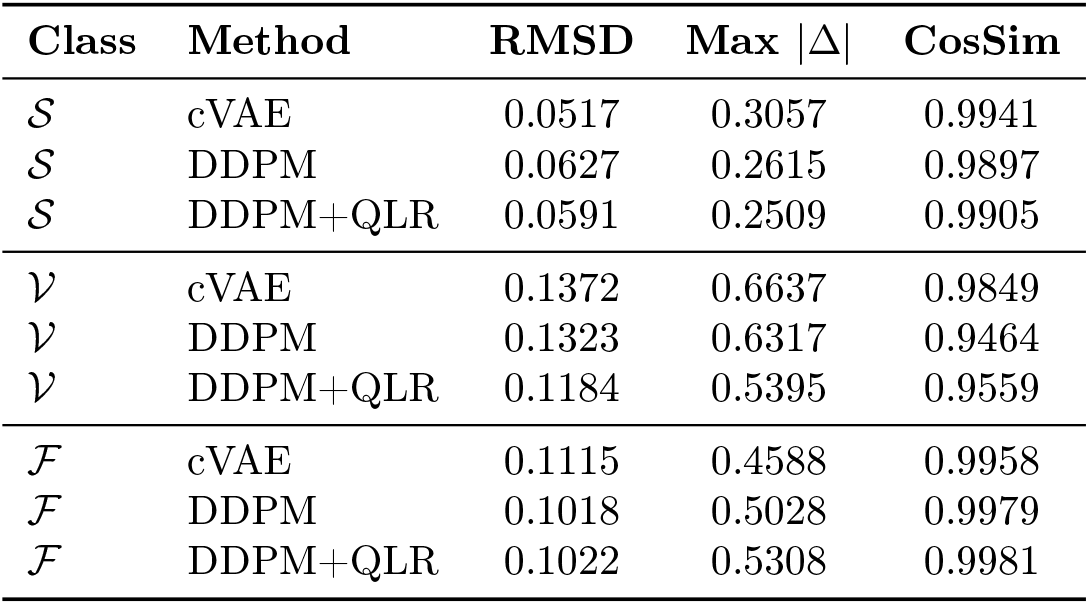
Morphological fidelity metrics comparing real and synthetic class-mean waveforms. RMSD: root-mean-square deviation. Max |Δ| : maximum point-wise absolute deviation. CosSim: cosine similarity. Lower RMSD and Max |Δ |and higher CosSim indicate better waveform alignment.

Table 2 summarizes the waveform-level fidelity metrics across all methods and minority classes. Differences between methods are small for all three classes, confirming that the primary synthesis quality is established at the cVAE and DDPM stages. The QLR module contributes most noticeably for Class 𝒱, the class of greatest clinical significance.

#### 3.1.3 Latent Space Alignment

Figure 15 shows PCA-projected latent distributions for the three minority classes, overlaying DDPM-generated samples against the kernel density estimate (KDE) of the real latent bank. The first two principal components explain 14.7% and 12.9% of the total latent variance, respectively.

**Figure 15:**
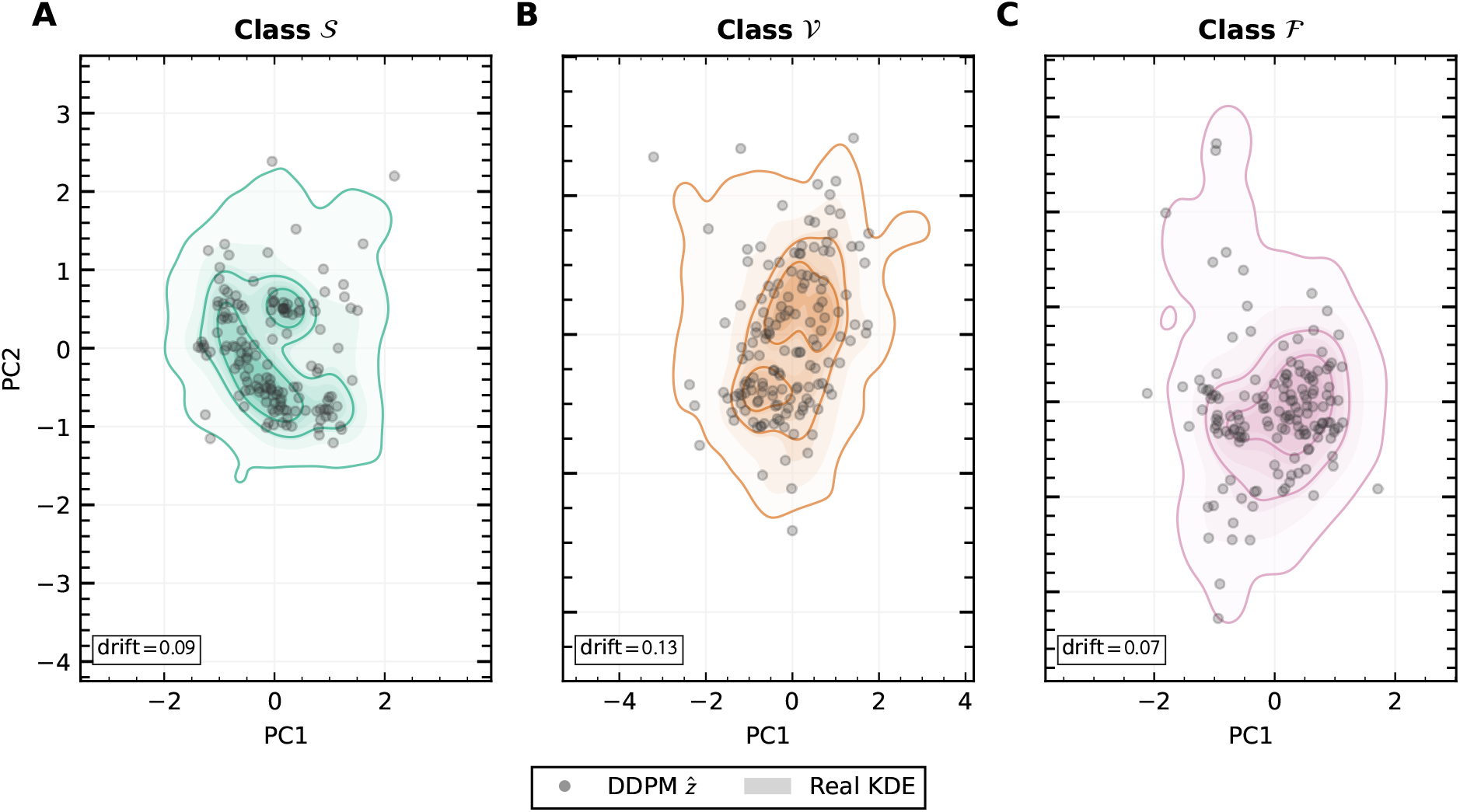
PCA projections of the 32-dimensional cVAE latent space for minority classes 𝒮 (A), 𝒱 (B), and ℱ (C). Grey scatter points are DDPM-generated latent samples, and shaded regions show the KDE of the real latent bank. Distribution drift values are 0.09, 0.13, and 0.07 for classes 𝒮, 𝒱, and ℱ, respectively.

To quantify how well the synthetic latent cloud covers the real class manifold, distribution drift is measured as the squared Maximum Mean Discrepancy between the real latent distribution *P* and the DDPM-generated distribution *Q*. Unlike the adaptive-bandwidth kernel used during QLR training (Section 2.5), this evaluation metric uses a mixture of *L* = 5 RBF kernels with fixed bandwidths *γ*_ℓ_ ∈ *{*0.5, 1.0, 2.0, 5.0, 10.0*}* to improve robustness against scale differences across classes. The per-kernel MMD^2^ values are averaged:

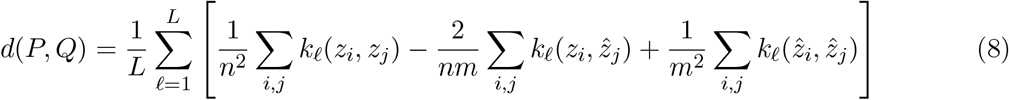

where *k*_ℓ_(*z, z*^*′*^) = exp −*γ*_ℓ_ ∥*z* − *z*^*′*^∥^2^ and *n, m* are the number of real and synthetic latent samples, respectively. A value of *d*(*P, Q*) = 0 indicates identical distributions. Larger values reflect systematic displacement of the synthetic cloud from the real manifold.

DDPM-generated samples broadly cover the support of each real distribution; distribution drift values of 0.09 (𝒮), 0.13 (𝒱), and 0.07 (ℱ) confirm reasonable fidelity across all three classes. Class 𝒱 exhibits the largest drift, consistent with the higher morphological complexity and broader intra-class variability of Ventricular ectopic beats.

Figures 16–18 illustrate the QLR-induced latent shifts for each minority class. Refined points (filled circles) consistently move toward higher-density regions of the real KDE relative to their DDPM origins (hollow circles), without collapsing to a single mode. This behavior confirms that the stay-loss and MMD objectives act in concert as intended: the stay-loss preserves distributional diversity and prevents degenerate solutions, while the MMD term pulls the synthetic cloud toward the real class manifold. Having established that the generative pipeline produces latent representations of high distributional fidelity, we now evaluate whether this latent-space quality translates into downstream classification gains.

**Figure 16:**
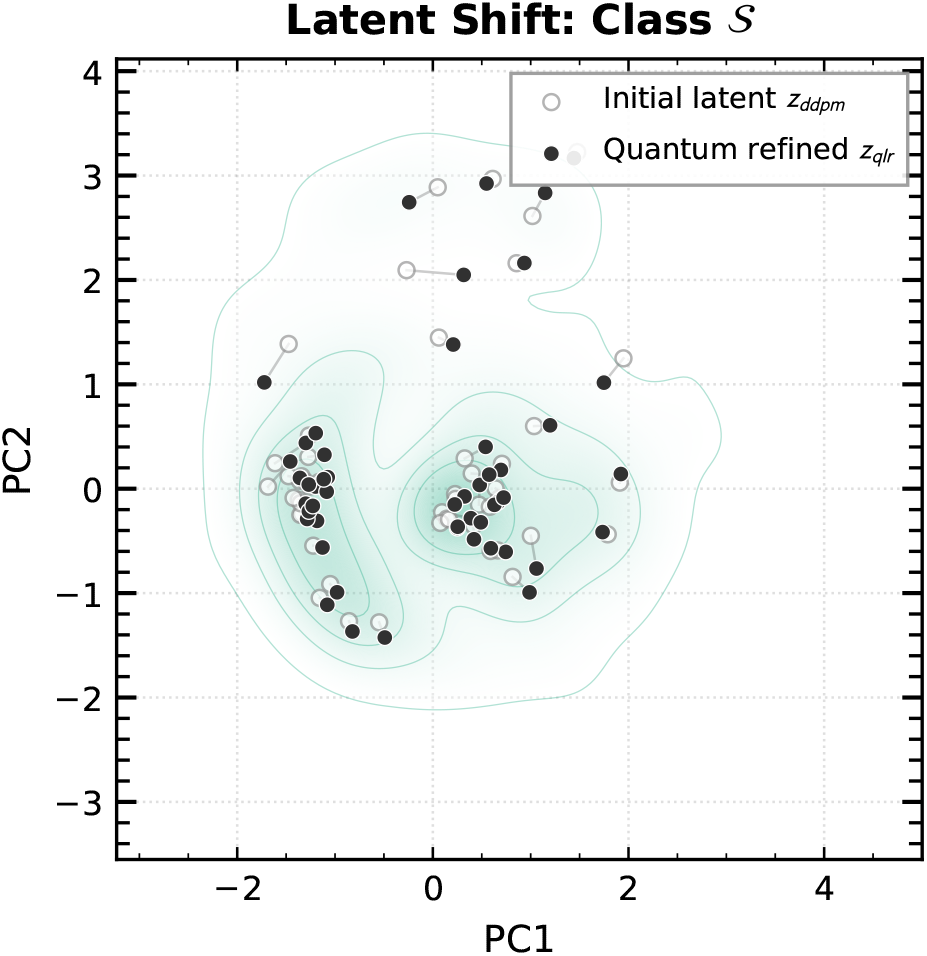
Latent shift induced by the QLR module for Class 𝒮. Hollow circles: DDPM latents before refinement; filled circles: quantum-refined latents, overlaid on the real latent KDE. Refined points shift toward higher-density regions without collapsing to a single mode.

**Figure 17:**
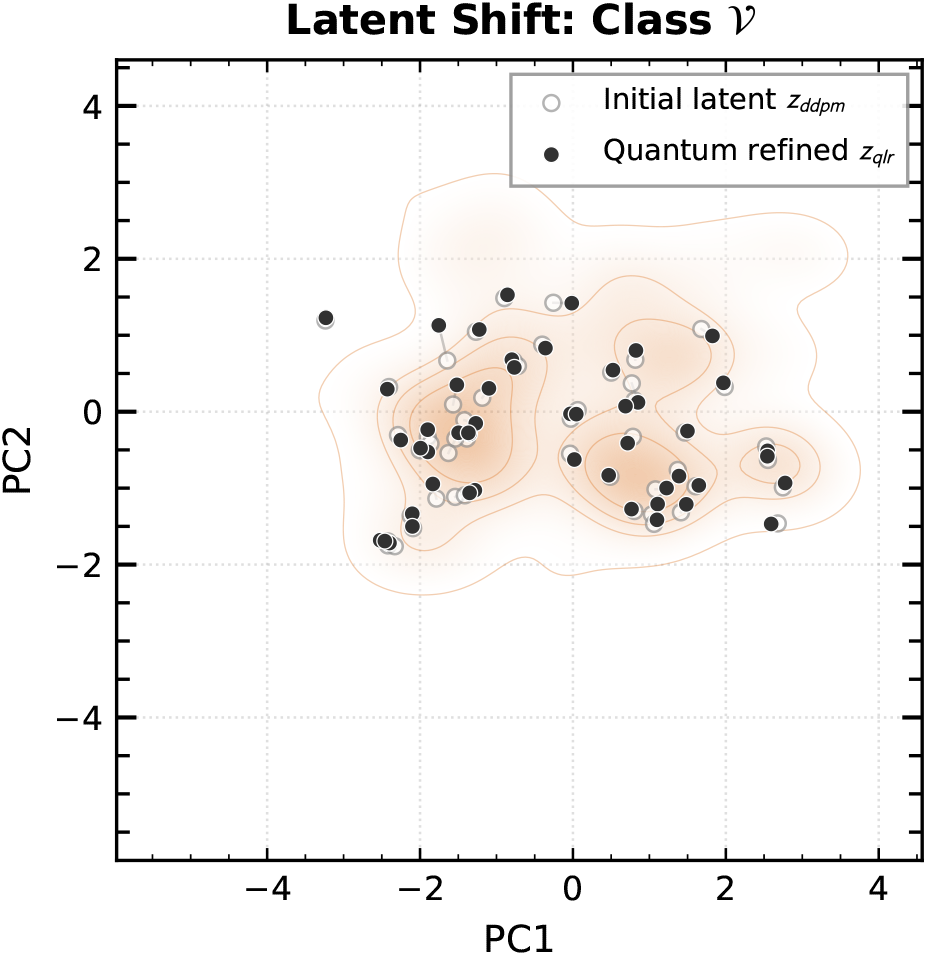
Latent shift induced by the QLR module for Class 𝒱. The QLR moves several outlier DDPM points toward the main density mass of the Ventricular latent distribution. The bounded correction (*α* = 0.10) prevents large displacements that would distort beat morphology.

**Figure 18:**
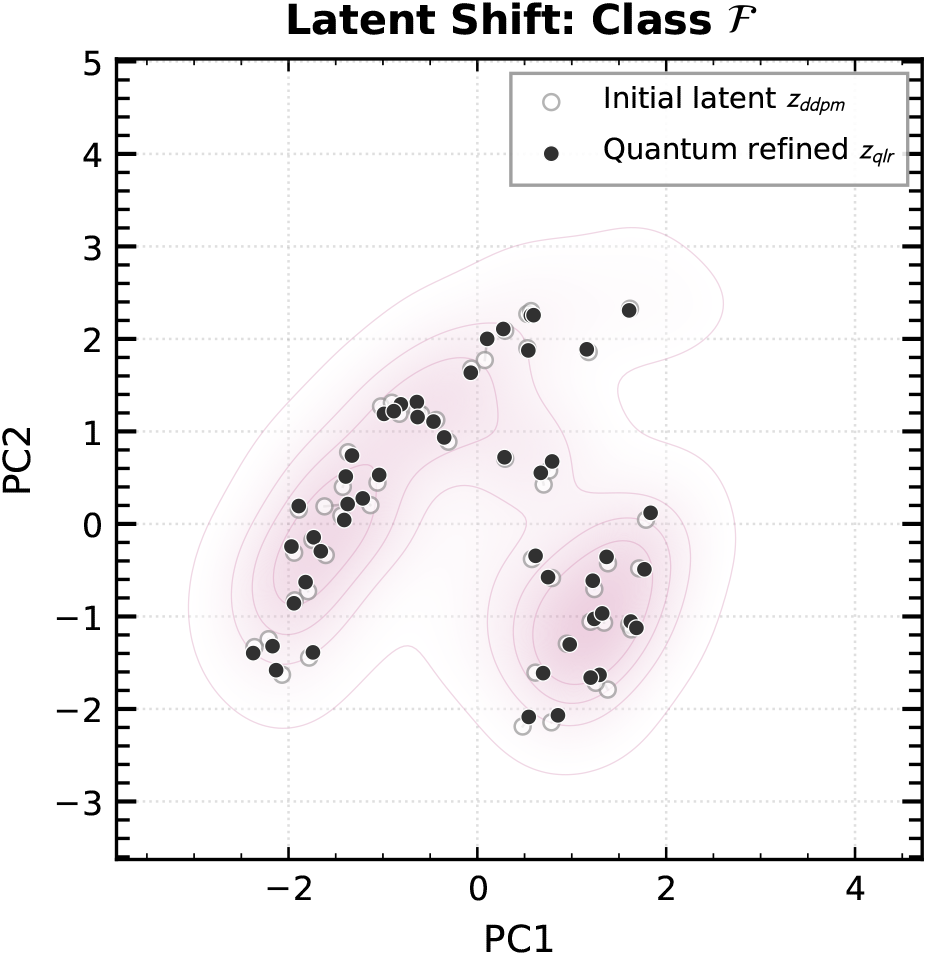
Latent shift induced by the QLR module for Class ℱ. The multi-modal Fusion latent distribution has two to three distinct clusters. The QLR realigns samples within each cluster without merging distinct modes, preserving intra-class morphological diversity.

### 3.2 Classification Performance

#### 3.2.1 Baseline and the Accuracy Paradox

The unaugmented classifier achieves a test accuracy of 0.943 ± 0.002 alongside a Macro F1 of only 0.747 ± 0.018 – a gap of nearly 20 percentage points. This is a direct manifestation of the accuracy paradox: the high accuracy is inflated by the ∼88.6% Normal-beat majority in the test set. This is consistent with the overall dataset distribution, since the intra-patient temporal split preserves each patient’s rhythm composition across the train and test windows – while per-class sensitivity for minority arrhythmias is substantially lower. In the best-seed baseline run, Class 𝒩 achieves 95.6% recall, but Class 𝒮 records only 60.9% recall, with 21.0% of Supraventricular beats misclassified as Ventricular. 𝒱-class recall reaches 97.3% but at the cost of only 67.9% precision, meaning that a large fraction of Normal beats are incorrectly flagged as Ventricular. This is an error rate that would generate clinically unacceptable false alarm volumes in a monitoring system (Figure 19).

**Figure 19:**
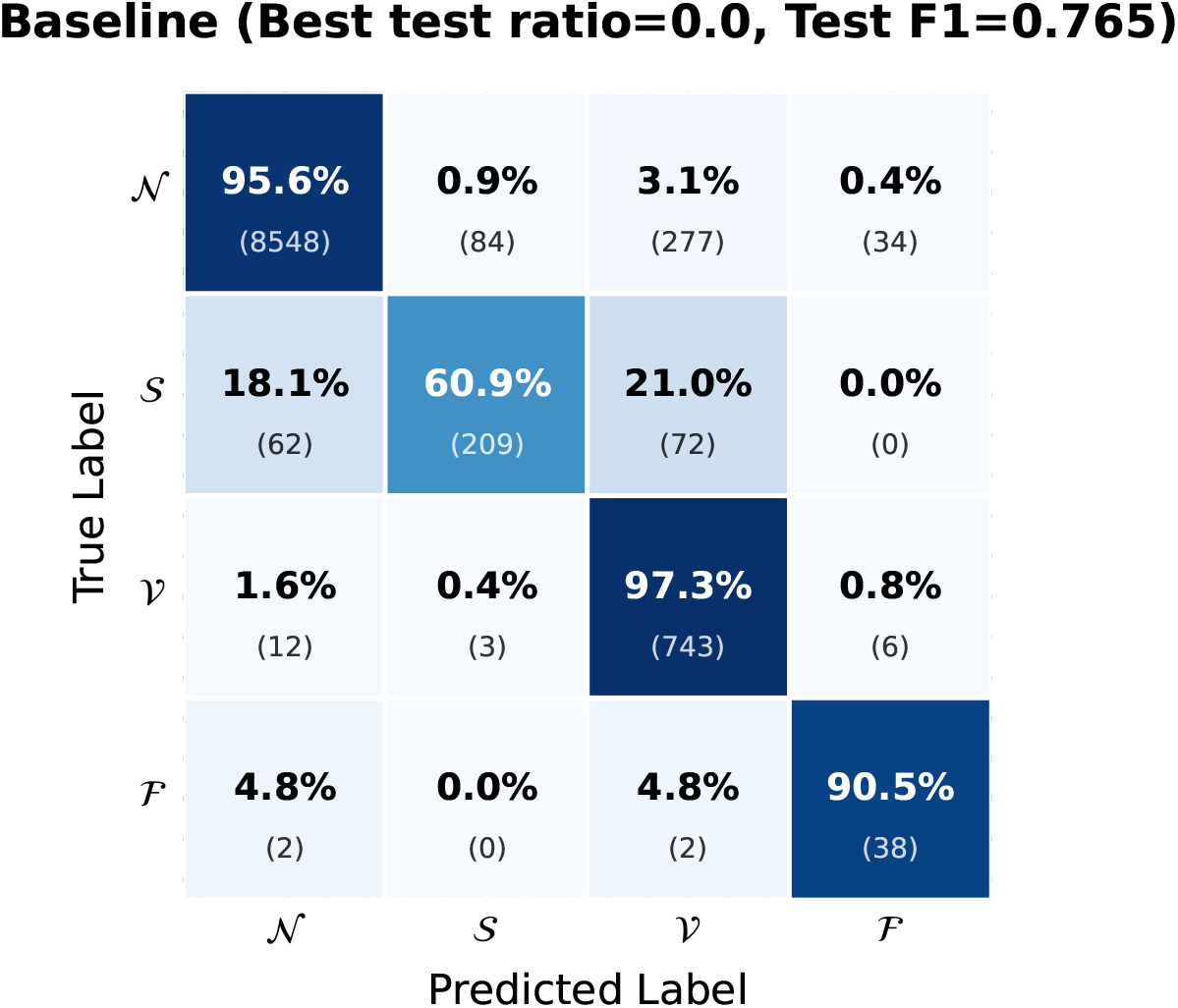
Confusion matrix for the unaugmented baseline (Macro F1 = 0.765). 𝒱-class recall is high (97.3%) but precision is only 67.9%, generating a high false Ventricular alarm rate. 𝒮-class recall is 60.9%, with 21.0% of Supraventricular beats misclassified as Ventricular.

The following subsections show how generative augmentation systematically addresses both the false alarm rate and the per-class sensitivity deficits observed in this baseline, and quantify the contribution of each pipeline stage.

#### 3.2.2 Overview of Augmentation Results

Table 3 reports mean Macro F1 and standard deviation over five independent random seeds for each method and ratio. All augmentation methods substantially improve over baseline. SMOTE provides consistent gains at low augmentation ratios, but its mean performance saturates and slightly degrades at *ρ* ≥ 0.75. All three deep generative methods (cVAE, DDPM, DDPM+QLR) outperform SMOTE at *ρ* = 0.75 and *ρ* = 1.00. DDPM+QLR achieves the highest mean Macro F1 of 0.835 ± 0.009 at *ρ* = 1.00.

**Table 3:** Macro F1 (mean ± std, five seeds) for each augmentation method across ratios *ρ* ∈ {0.25, 0.50, 0.75, 1.00}. Unaugmented baseline: 0.747 ± 0.018. Best result per ratio shown in bold.

| Method | $\rho = 0.25$ | $\rho = 0.50$ | $\rho = 0.75$ | $\rho = 1.00$ |
| --- | --- | --- | --- | --- |
| SMOTE | $0.811 \pm 0.025$ | $0.813 \pm 0.014$ | $0.802 \pm 0.024$ | $0.803 \pm 0.017$ |
| cVAE | **$0.821 \pm 0.017$** | **$0.831 \pm 0.013$** | **$0.832 \pm 0.014$** | $0.830 \pm 0.014$ |
| DDPM | $0.815 \pm 0.025$ | $0.816 \pm 0.011$ | $0.820 \pm 0.012$ | $0.826 \pm 0.016$ |
| DDPM+QLR | $0.809 \pm 0.013$ | $0.815 \pm 0.025$ | $0.828 \pm 0.013$ | **$0.835 \pm 0.009$** |

Table 4 provides a complete per-class breakdown of precision and recall for each method at its best augmentation ratio (SMOTE: *ρ* = 0.50, cVAE: *ρ* = 0.75, DDPM and DDPM+QLR: *ρ* = 1.00). Each cell reports the mean and standard deviation over five random seeds. The table reveals several patterns that are not visible in the aggregate Macro F1. Macro-averaged precision rises consistently from 0.698 ± 0.019 at baseline to 0.859 ± 0.015 under DDPM+QLR, while macro-averaged recall changes more modestly, from 0.839 ± 0.013 to 0.827 ± 0.018. This reflects the fact that augmentation primarily corrects precision deficits rather than uniformly boosting recall across all classes. The dominant driver of precision improvement is the 𝒱 class, which rises from 0.692 ± 0.025 at baseline to 0.864 ± 0.078 under DDPM+QLR. This is a reduction in false Ventricular alarms that directly translates into clinical viability. 𝒮-class recall remains near 60% across all methods, confirming that augmentation addresses the false-alarm problem more effectively than the missed-detection problem for Supraventricular beats. ℱ-class results carry the highest variance across all methods, consistent with the very small test set of 42 Fusion beats.

**Table 4:** Per-class precision (PPV) and recall (SEN) for each method at its best augmentation ratio (mean ± std, five seeds). Each augmented method is evaluated at the ratio maximizing its mean Macro F1 (SMOTE: *ρ* = 0.50, cVAE: *ρ* = 0.75, DDPM and DDPM+QLR: *ρ* = 1.00). Best value per metric shown in bold. Top: macro-averaged PPV and SEN across all classes, and Classes 𝒩 and 𝒮. Bottom: Classes 𝒱 and ℱ.

| Method | All Classes | | Class $\mathcal{N}$ | | Class $\mathcal{S}$ | |
| --- | --- | --- | --- | --- | --- | --- |
|  | PPV | SEN | PPV | SEN | PPV | SEN |
| Baseline | $0.698 \pm 0.019$ | **$0.839 \pm 0.013$** | $0.989 \pm 0.005$ | $0.954 \pm 0.002$ | $0.667 \pm 0.035$ | **$0.598 \pm 0.007$** |
| SMOTE | $0.820 \pm 0.022$ | $0.822 \pm 0.007$ | **$0.992 \pm 0.003$** | $0.983 \pm 0.005$ | $0.818 \pm 0.040$ | $0.597 \pm 0.018$ |
| cVAE | $0.853 \pm 0.036$ | $0.828 \pm 0.010$ | $0.984 \pm 0.001$ | $0.983 \pm 0.013$ | $0.841 \pm 0.055$ | $0.576 \pm 0.023$ |
| DDPM | $0.857 \pm 0.031$ | $0.819 \pm 0.011$ | $0.989 \pm 0.005$ | $0.984 \pm 0.009$ | **$0.912 \pm 0.021$** | $0.579 \pm 0.017$ |
| DDPM+QLR | **$0.859 \pm 0.015$** | $0.827 \pm 0.018$ | $0.984 \pm 0.002$ | **$0.985 \pm 0.008$** | $0.860 \pm 0.028$ | $0.591 \pm 0.017$ |

| Method | Class $\mathcal{V}$ | | Class $\mathcal{F}$ | |
| --- | --- | --- | --- | --- |
|  | PPV | SEN | PPV | SEN |
| Baseline | $0.692 \pm 0.025$ | $0.974 \pm 0.003$ | $0.446 \pm 0.085$ | **$0.829 \pm 0.046$** |
| SMOTE | $0.802 \pm 0.053$ | **$0.974 \pm 0.005$** | $0.668 \pm 0.055$ | $0.733 \pm 0.046$ |
| cVAE | $0.848 \pm 0.086$ | $0.968 \pm 0.011$ | **$0.738 \pm 0.041$** | $0.786 \pm 0.038$ |
| DDPM | $0.804 \pm 0.096$ | $0.971 \pm 0.004$ | $0.722 \pm 0.043$ | $0.743 \pm 0.062$ |
| DDPM+QLR | **$0.864 \pm 0.078$** | $0.961 \pm 0.015$ | $0.730 \pm 0.036$ | $0.771 \pm 0.073$ |

Figures 20–23 show the confusion matrices for the seed 42 classifiers of each augmentation method at their best augmentation ratio. These are presented as representative examples. All quantitative conclusions are drawn from the five-seed means reported in Table 3 and Table 4. All methods substantially reduce the 𝒩 -class false positive rate, improving 𝒩 -class recall from 95.6% (baseline) to approximately 98–99%. The primary mechanism behind the Macro F1 improvement is not a uniform increase in all class-wise recalls. Rather, it is a dramatic increase in 𝒱-class precision, driven by the reduction in Normal beats being misclassified as Ventricular.

**Figure 20:**
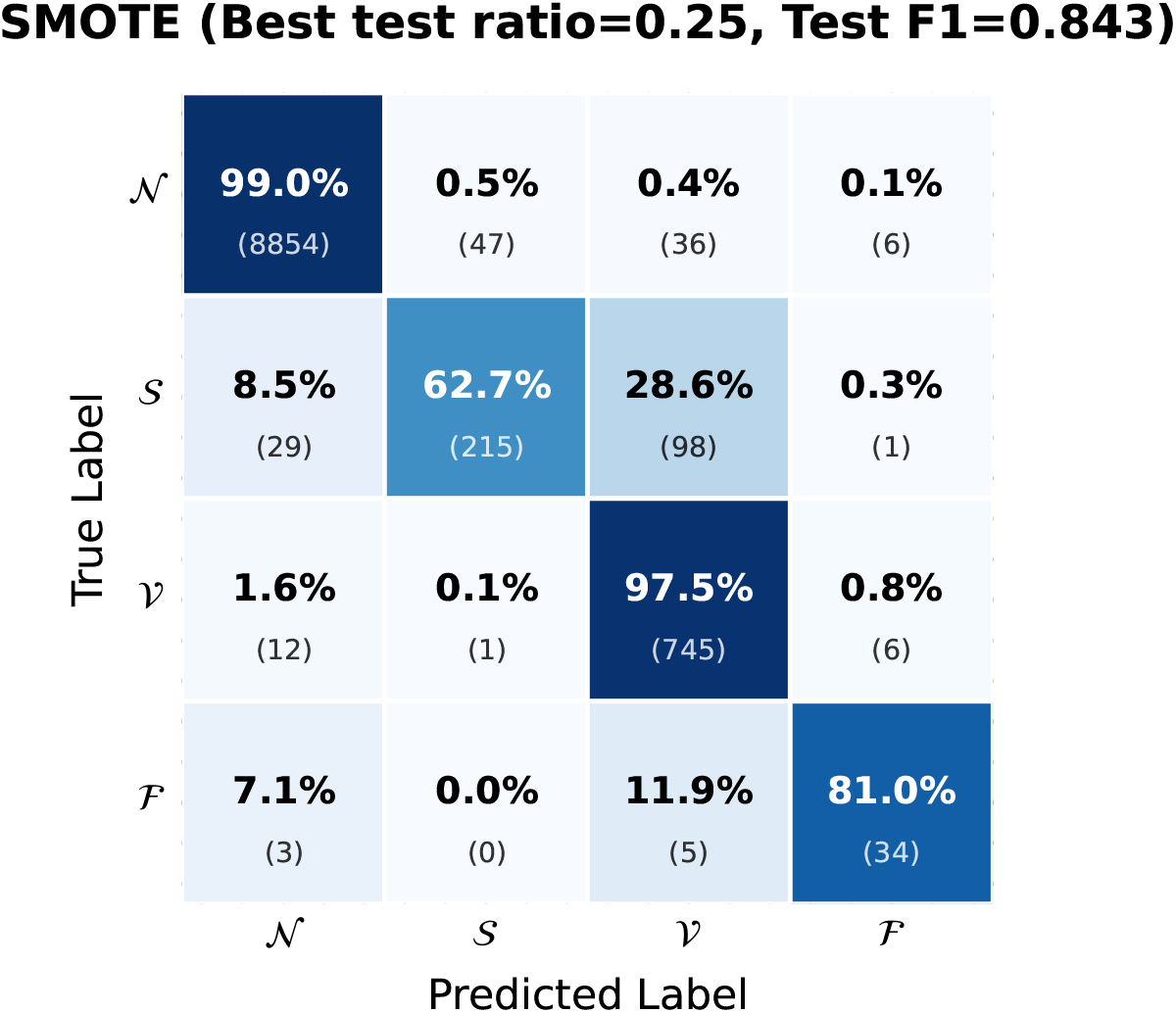
SMOTE (*ρ* = 0.25). ℱ-class recall reaches 81.0%, but 28.6% of -class beats are misclassified as Ventricular—considerably higher than for the deep generative methods—suggesting that SMOTE-interpolated 𝒮 beats overlap with the Ventricular feature space.

**Figure 21:**
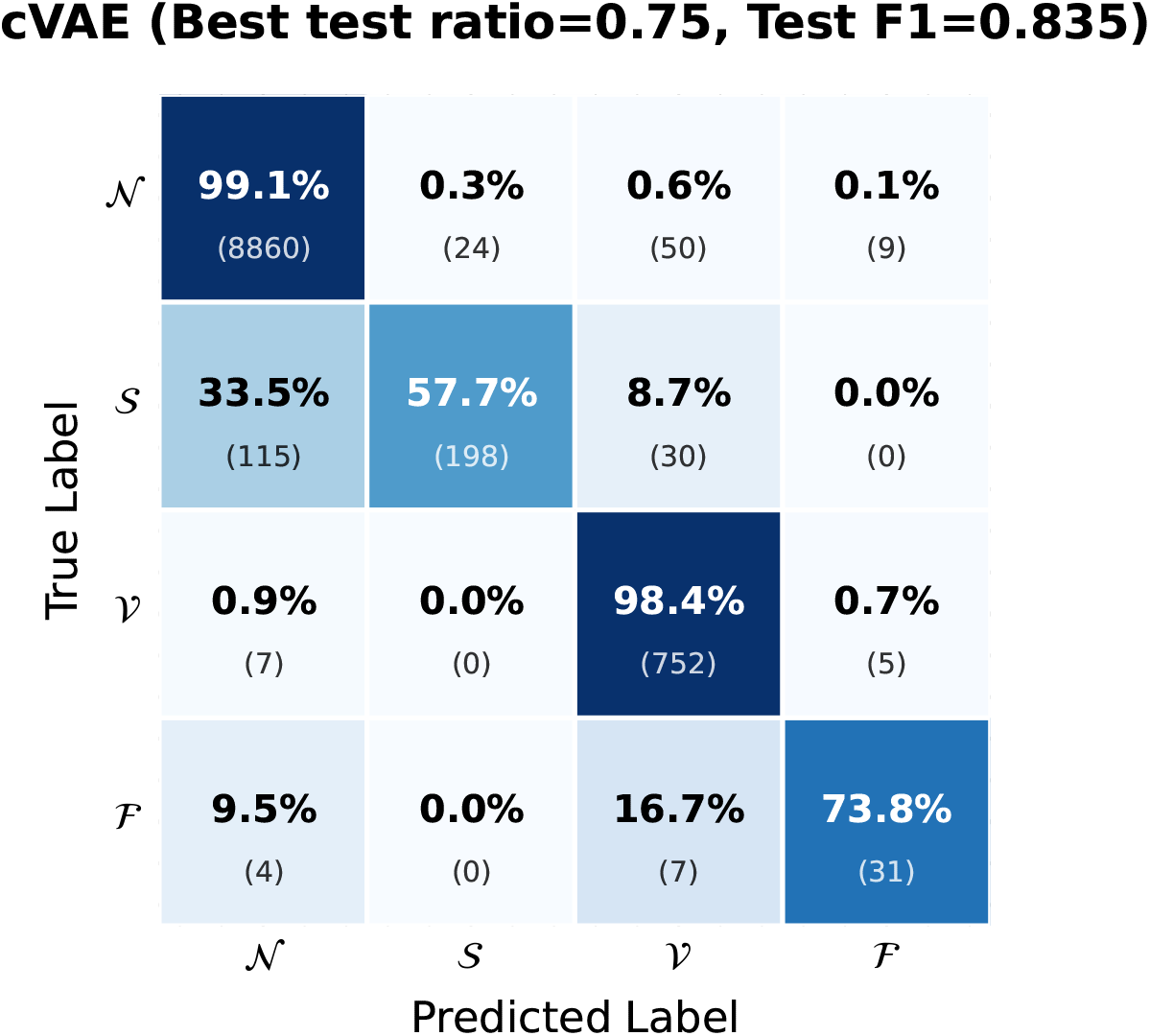
cVAE (*ρ* = 0.75). 𝒩-class recall improves to 99.1% and 𝒱-class precision improves markedly over baseline.

**Figure 22:**
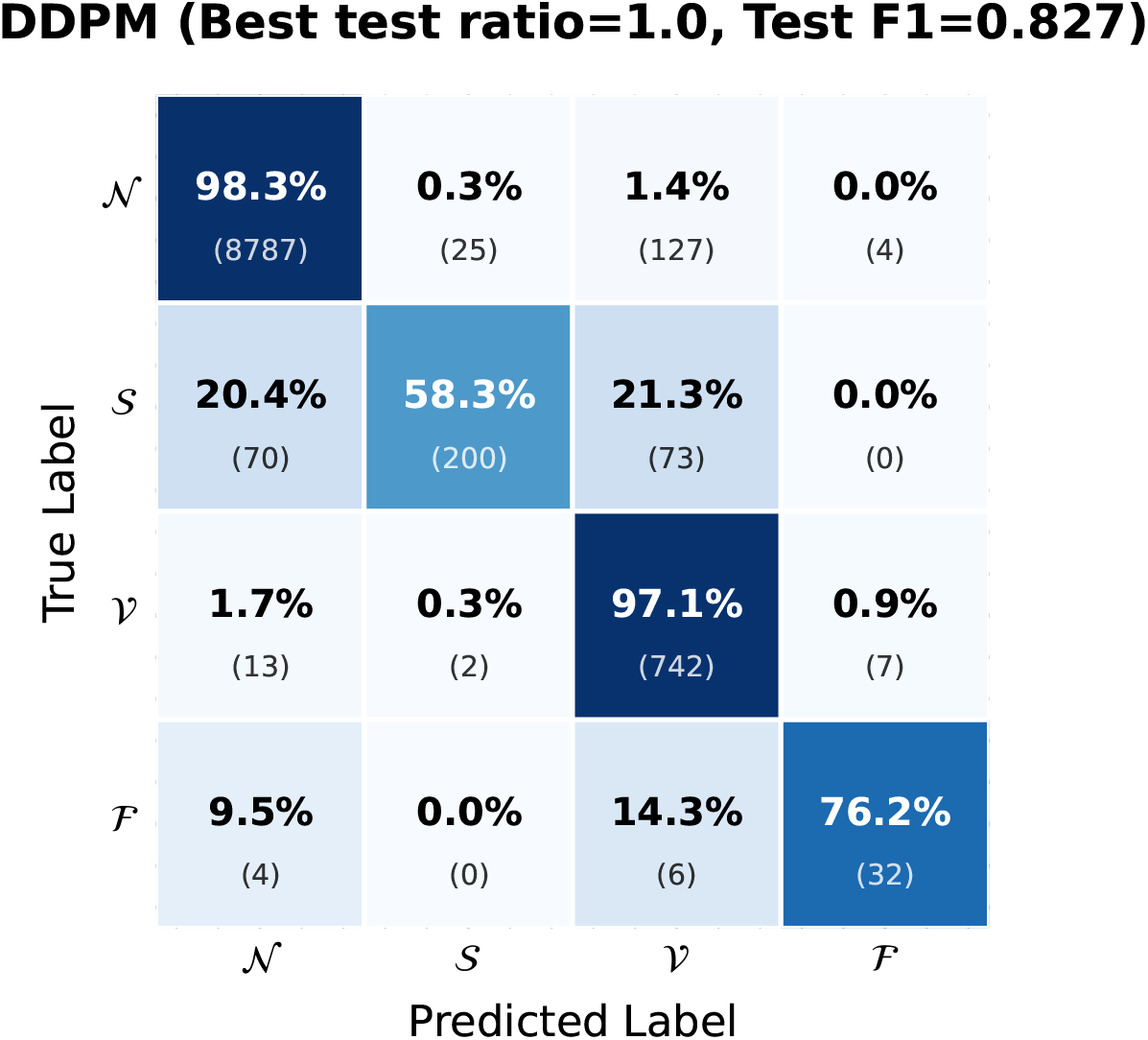
DDPM (*ρ* = 1.00). 𝒩 -class recall rises to 98.3%, 𝒱-class precision improves substantially, and 𝒮→ 𝒱 confusion falls to 8.7%.

**Figure 23:**
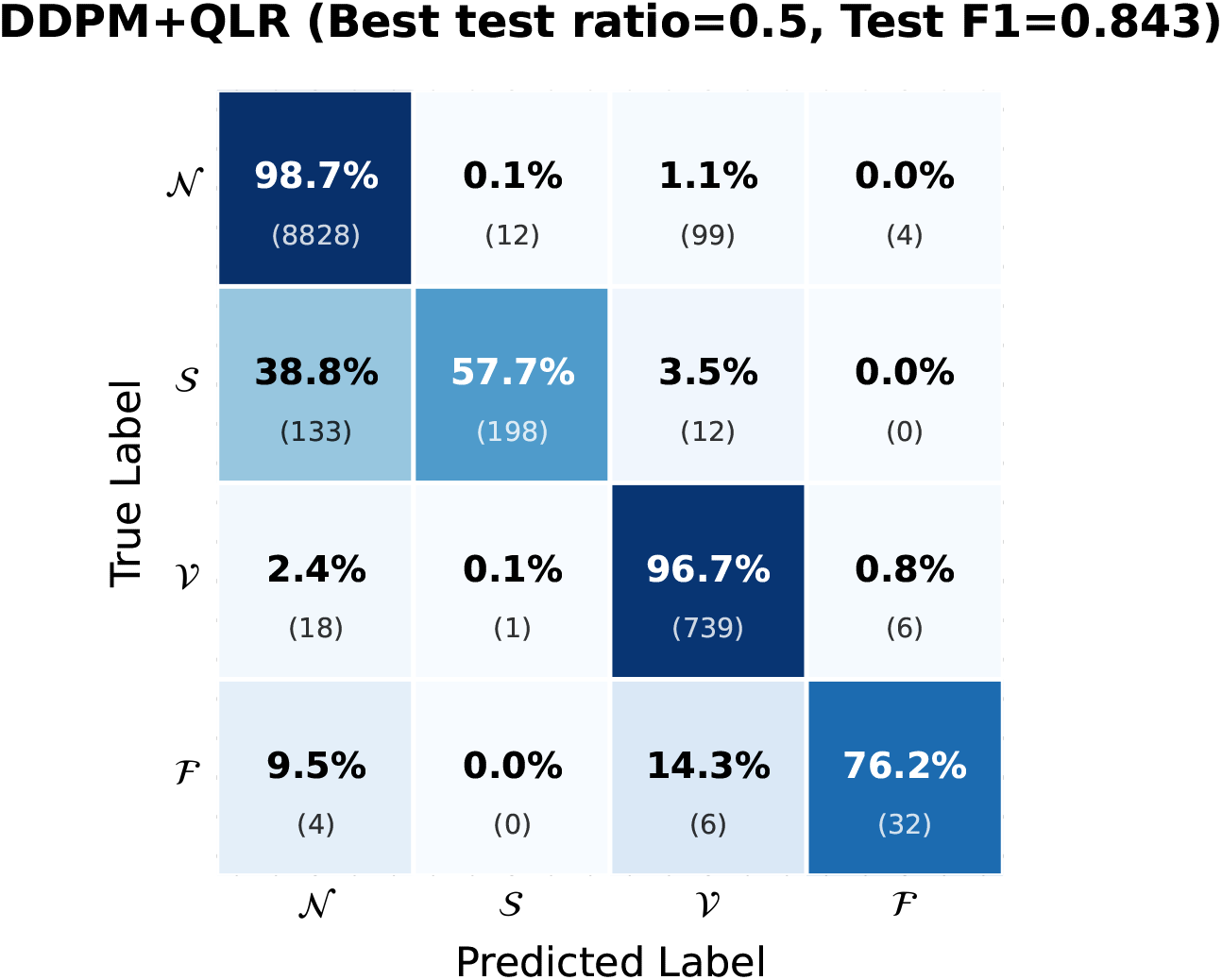
DDPM+QLR (*ρ* = 0.50). 𝒱-class precision reaches 86.3% and 𝒱-class F1 reaches ∼0.91. 𝒮→𝒱 confusion drops to 3.5%, compared to 28.6% under SMOTE.

#### 3.2.3 Ensemble Inference

A five-model soft-voting ensemble combining the DDPM+QLR classifiers trained at *ρ* = 1.00 across all five seeds achieves a test Macro F1 of 0.856 – the highest overall result in this study. The ensemble operates globally. Each of the five models produces a four-class probability vector for every test beat, and the vectors are averaged before taking the argmax. No patient-specific model selection or per-record weighting is applied. Class-wise results are: 𝒱 -class F1 = 0.987, 𝒮-class F1 = 0.733, 𝒱-class F1 = 0.929, ℱ-class F1 = 0.777, weighted-average F1 = 0.973. The ensemble 𝒱-class result (recall 97.4%, precision 88.8%) represents a substantially improved operating point compared to the baseline precision of 67.9%, reducing the false Ventricular alarm rate to a clinically more acceptable level. The 𝒮-class and ℱ-class ensemble F1 values also improve relative to single-seed means, as soft-voting effectively pools complementary decision boundaries from different random initializations.

### 3.3 Minority Class Analysis and Ablation

𝒮-class recall remains between 57% and 61% across all methods and seeds and does not improve meaningfully with augmentation. The ensemble 𝒮-class F1 of 0.733 (versus baseline ∼0.65) is driven primarily by precision: in the best DDPM+QLR run, 𝒮-class precision reaches 93.8% compared to approximately 70.6% at baseline, indicating that augmentation sharply reduces false Supraventricular predictions without substantially increasing true positive recovery.

The ℱ class, with only 708 training beats and 42 test beats, has highly variable single-seed performance estimates. The ensemble ℱ-class F1 of 0.777 is the most stable ℱ-class result obtained and is clinically encouraging, although robust conclusions for this class would require a dataset with substantially more Fusion beat instances.

The ablation is conducted on Class 𝒱 (seed 42), which is the most data-rich minority class (5,527 training beats) and therefore provides the most statistically stable MMD estimates for comparing the two refiners. 𝒮 and ℱ class ablations are feasible but less informative at this sample scale. The small real-beat pools (2,186 and 708 beats respectively) produce high-variance MMD estimates that would make it difficult to attribute any observed difference between the two refiners to a systematic effect rather than sampling noise. The QLR and a matched classical Multi-Layer Perceptron (MLP) refiner of equivalent parameter count were trained for 80 epochs under identical optimization conditions. At convergence, the MLP reaches a lower final validation loss than the QLR (0.0038 vs. 0.0048), consistent with the expectation that an unconstrained classical approximator optimizes a scalar objective more efficiently than a quantum circuit operating through expectation-value readout. The QLR is not intended as an optimizer but as a distributional regularizer, and the downstream classification gain of DDPM+QLR over plain DDPM remains directional and consistent across all augmentation ratios despite the higher validation loss.

The PQC offers two properties that a classical MLP of equivalent parameter count does not. First, the quantum feature map embeds the 32-dimensional latent input into a 2^8^ = 256-dimensional Hilbert space, providing access to a richer class of inter-dimensional correlations per parameter than a classical network of the same size.^47^ Second, the layered rotation – entanglement architecture imposes a structured inductive bias over latent co-variation that differs in character from the isotropic smoothing of a fully connected MLP. These properties suggest that the PQC’s contribution is best understood as Hilbert-space regularization: it does not find a better minimum, but it constrains the correction to a structured manifold that happens to be better matched to the distributional correction task. The paired *t*-test (*p* = 0.407) confirms that the gain is directional but not statistically significant under the five-seed protocol. A powered comparison would require either more seeds or a larger dataset. This is the first application of a PQC as a latent-space distributional refiner within a generative augmentation pipeline, distinct from prior work employing quantum circuits as direct classifiers,^25^ generators,^29^ anomaly detectors,^30^ or latent diffusion models.^31^

## 4 Discussion

The unaugmented baseline illustrates the accuracy paradox directly and explains why augmentation is necessary. A classifier with 94.3% test accuracy and only 74.7% Macro F1 is largely unsuited for detecting rare but clinically significant arrhythmias; any evaluation framework reporting only accuracy in this setting would systematically overstate the system’s diagnostic utility.^10^ The accuracy paradox documented here is not an artifact of model choice but a direct consequence of training on a severely imbalanced distribution, and the systematic Macro F1 improvements produced by all generative methods confirm that it is correctable through principled augmentation.

The generative pipeline produces synthetic minority-class beats with high morphological fidelity – most consequentially for Class 𝒱 – as confirmed by the waveform-level metrics in Table 2. Both the cVAE and DDPM stages independently establish strong reconstruction quality, capturing the dominant morphological signatures of each arrhythmia class. The most clinically relevant morphological finding is the QLR module’s contribution to Class 𝒱. The PQC’s bounded latent correction pulls outlier DDPM samples toward the high-density regions of the Ventricular latent manifold, reducing both RMSD and maximum absolute waveform deviation. This is precisely the class that matters most from a prognostic standpoint. Frequent Ventricular ectopy – particularly complex forms such as bigeminy, couplets, or non-sustained runs – can precede life-threatening ventricular arrhythmias and may warrant pharmacological or procedural intervention, so both the sensitivity and precision of Ventricular detection carry direct clinical weight. The ensemble 𝒱-class operating point of 97.4% recall and 88.8% precision is clinically viable for wearable Holter-style monitoring: nearly all true Ventricular events are correctly identified, and the false positive rate is low enough to avoid the alert fatigue that undermines clinical trust in automated arrhythmia monitoring systems. The most practically significant single result in this work is therefore the improvement in 𝒱-class precision from 67.9% at baseline to 88.8% in the DDPM+QLR ensemble –a change that would meaningfully reduce the burden of false alerts on clinicians and patients.

Supraventricular ectopic beats present a different and more persistent challenge than Ventricular ectopy, revealing a ceiling on 𝒮-class recall that warrants careful clinical interpretation. 𝒮-class recall does not exceed 60% for any single-seed run across all augmentation methods and all ratios. This ceiling likely reflects the intrinsic morphological proximity between Supraventricular ectopic beats and certain Normal beat variants under the intra-patient temporal split: if a patient’s sinus rhythm morphology evolves between the calibration and test windows – due to rate-related aberrancy or medication effects for example – the 𝒮/ 𝒩 boundary in feature space shifts in a way that augmentation cannot compensate for. The improvement in 𝒮-class precision rather than recall suggests that augmentation primarily makes the classifier more conservative about assigning the 𝒮 label, reducing one class of diagnostic error while leaving another largely unaddressed. Improving 𝒮-class recall without sacrificing 𝒩 -class specificity likely requires approaches beyond static data augmentation, such as patient-specific temporal context modeling or explicit morphological feature engineering targeting P-wave characteristics.

SMOTE’s plateau and degradation at high augmentation ratios align with the well-documented limitations of linear interpolation in high-dimensional biomedical feature spaces,^16^ and explain why deep generative methods are preferable at large synthesis volumes. When a large volume of interpolated beats fills the training set, the natural cluster structure of minority distributions is smoothed over, and boundary-straddling samples add noise to the decision surface. This is most strikingly illustrated by the confusion matrix for the seed 42 SMOTE classifier (Figure 20): 28.6% of 𝒮-class beats are misclassified as Ventricular, compared to only 3.5% under DDPM+QLR. The elevated 𝒮→ 𝒱 confusion rate under SMOTE is a concrete clinical consequence: SMOTE generates Supraventricular beats that encroach on the Ventricular feature space, creating a clinically dangerous misclassification pattern in which 𝒮-class beats are assigned the more alarming Ventricular label. The deep generative methods avoid this by modeling the underlying distribution directly rather than interpolating between observed points, and their 𝒮→ 𝒱 confusion rates are substantially lower as a result.

The ablation study complicates a straightforward quantum-advantage interpretation, and suggests that the QLR’s role is best understood as distributional regularization rather than raw optimization. The PQC imposes a structured inductive bias through its quantum feature map: the layered rotation–entanglement architecture introduces correlations across latent dimensions that differ from those induced by a classical MLP with the same parameter count. This bias appears to be better suited to the distributional correction task as measured by downstream classification quality, even if not by validation loss. The QLR’s contribution is therefore best understood as a form of Hilbert-space regularization rather than a demonstration of quantum computational superiority in the traditional optimization sense.^21^ The non-significant *p*-value under the five-seed protocol means this interpretation cannot be fully confirmed from the available data. A statistically powered comparison would require a larger number of seeds or a larger dataset.

Several important limitations apply to this work. The quantum circuit is evaluated entirely on the PennyLane^45^ `default.qubit` classical simulator, which does not model hardware-level noise sources such as qubit decoherence, gate fidelity errors, or readout noise. Performance on actual near-term quantum hardware would likely be lower, and the results presented here should be interpreted as a proof of principle for the architectural design rather than a hardware-ready deployment. The intra-patient temporal split answers the personalized monitoring question but does not establish inter-patient generalizability. A patient-calibrated model cannot be assumed to generalize to a different patient without retraining. Additionally, the ℱ-class test set of 42 beats is too small for statistically reliable single-seed evaluation, and Fusion-beat results should be treated with cor esponding caution. Finally, the QLR module trains a separate quantum circuit per minority class, making the quantum simulation stage the most computationally expensive component of the pipeline and limiting its practical scalability in its current form.

## 5 Conclusions

This study introduces and evaluates a three-stage hybrid quantum-classical generative pipeline for imbalanced ECG arrhythmia classification. A spectral-guided cVAE compresses beat-level ECG signals into a stable 32-dimensional latent space; a class-conditional latent DDPM generates new minority-class latents; and a Quantum Latent Refinement module built on an 8-qubit PQC applies a bounded MMD-guided distributional correction to align synthetic latents with the real class-specific latent manifold. Downstream classification is performed by a lightweight 1D MobileNetV2 evaluated across five seeds and four augmentation ratios.

All generative augmentation methods substantially outperform the unaugmented baseline, improving Macro F1 from 0.747 ± 0.018 to a range of 0.820–0.835 at peak ratios. DDPM+QLR achieves the highest mean Macro F1 at *ρ* = 1.00, and a five-model soft-voting ensemble reaches 0.856. The QLR gain over plain DDPM is directional and consistent across augmentation ratios but does not reach statistical significance under the five-seed protocol, framing the quantum module as a distributional regularizer rather than a raw optimization advantage. Across all methods, augmentation primarily corrects precision deficits rather than uniformly boosting recall: macro-averaged precision improves from 0.698 ± 0.019 at baseline to 0.859 ± 0.015 under DDPM+QLR, while macro-averaged recall changes more modestly from 0.839 ± 0.013 to 0.827 ± 0.018, reflecting the asymmetric nature of the class imbalance problem. The most clinically actionable result is the ensemble 𝒱-class performance (recall 97.4%, precision 88.8%), which represents a substantial and clinically meaningful improvement over the baseline’s false positive rate and offers a viable operating point for wearable Holter-style arrhythmia monitoring.

Future directions include hardware-aware PQC designs tolerant of realistic noise models, and extension of the QLR ablation to all minority classes with a larger number of seeds for statistically powered comparisons. Evaluating inter-patient generalization across multiple ECG databases and integrating end-to-end R-peak detection would move the pipeline toward fully deployable wearable monitoring.

## Data Availability

The MIT-BIH Arrhythmia Database is publicly available at https://physionet.org/content/mitdb/. Code for the generative pipeline and downstream classifier will be made publicly available on GitHub upon acceptance. In the meantime, code is available upon reasonable request from the corresponding author.

## Author Contributions

**G. Kritopoulos:** Conceptualization (equal); Formal analysis (equal); Investigation (equal); Methodology (equal); Software (equal); Visualization (equal); Writing – original draft preparation and revision (equal).

**G. Neofotistos:** Conceptualization (equal); Methodology (equal); Supervision (equal); Validation (equal); Writing – review and editing (equal).

**G. D. Barmparis:** Conceptualization (equal); Investigation (equal); Supervision (equal); Validation (equal); Writing – review & editing (equal).

**G. P. Tsironis:** Conceptualization (equal); Funding acquisition (equal); Investigation (equal); Supervision (equal); Validation (equal); Writing – review & editing (equal).

## Institutional Review Board Statement

This study used exclusively the publicly available, de-identified MIT-BIH Arrhythmia Database (PhysioNet; https://physionet.org/content/mitdb/), which was collected under informed consent at Beth Israel Hospital, Boston, MA, USA. No new human subjects were enrolled. Ethical review and approval were waived for this study, as it constitutes a secondary analysis of a fully de-identified, open-access dataset that does not allow re-identification of individual participants.

## Informed Consent Statement

Not applicable. This study used only the publicly available, de-identified MIT-BIH Arrhythmia Database. No new patient data were collected.

## Data Availability and Statistical Reporting

The MIT-BIH Arrhythmia Database is publicly available at https://physionet.org/content/mitdb/.^9^ Results are reported as mean ± standard deviation over five independent random seeds. Statistical comparisons between augmentation methods use a paired *t*-test (`scipy.stats. ttest_rel`) on seed-wise Macro F1 values. Code for the generative pipeline and downstream classifier will be made publicly available on GitHub upon acceptance. In the meantime, code is available upon reasonable request from the corresponding author.

